# Design and implementation of an international, multi-arm, multi-stage platform master protocol for trials of novel SARS-CoV-2 antiviral agents: Therapeutics for Inpatients with COVID-19 (TICO/ACTIV-3)

**DOI:** 10.1101/2020.11.08.20227876

**Authors:** Daniel D. Murray, Abdel G. Babiker, Jason V. Baker, Christina E. Barkauskas, Samuel M. Brown, Christina C. Chang, Victoria J. Davey, Annetine C. Gelijns, Adit A. Ginde, Birgit Grund, Elizabeth Higgs, Fleur Hudson, Virginia L. Kan, H. Clifford Lane, Thomas A. Murray, Roger Paredes, Mahesh K.B. Parmar, Sarah Pett, Andrew N. Phillips, Mark N. Polizzotto, Cavan Reilly, Uriel Sandkovsky, Shweta Sharma, Marc Teitelbaum, B. Taylor Thompson, Barnaby E. Young, James D. Neaton, Jens D. Lundgren, on behalf of the TICO Study Group

**Affiliations:** CHIP Centre of Excellence for Health, Immunity, and Infections, Department of Infectious Diseases, Rigshospitalet, Copenhagen, Denmark; Medical Research Council Clinical Trials Unit at UCL, University College London, London, UK; Hennepin Healthcare Research Institute, Minneapolis, Minnesota, USA; University of Minnesota, Minneapolis, MN, USA; Division of Pulmonary, Allergy, and Critical Care Medicine, Department of Medicine, Duke University, Durham, NC 27710, USA; Intermountain Medical Center, Murray, UT, USA; University of Utah School of Medicine, Salt Lake City, UT, USA; The Kirby Institute, University of New South Wales, Sydney, Australia; U.S. Department of Veterans Affairs, Washington, D.C., USA; Department of Population Health Science and Policy, Icahn School of Medicine at Mount Sinai, New York, NY, USA; Department of Emergency Medicine, University of Colorado School of Medicine, Aurora, CO, USA; Division of Biostatistics, School of Public Health, University of Minnesota, Minneapolis, MN, USA; National Institute of Allergy and Infectious Diseases, Bethesda, MD, USA; MRC Clinical Trials Unit at UCL, Institute of Clinical Trials & Methodology, London; Veteran Affairs Medical Center, Washington, D.C., USA; George Washington University School of Medicine and Health Sciences, Washington, D.C., USA; Infectious Diseases Department & irsiCaixa AIDS Research Institute, Hospital Universitari Germans Trias i Pujol, Catalonia, Spain; Institute for Global Health, University College London, London, UK; St Vincent’s Hospital, Sydney, Australia; Division of Infectious Diseases, Baylor University Medical Center, Dallas, TX, USA; Leidos Biomedical Research, Inc., Frederick, MD, USA; Division of Pulmonary and Critical Care, Department of Medicine, Massachusetts General Hospital, Boston, MA, USA; Harvard Medical School Boston, MA, USA; National Centre for Infectious Diseases, Singapore, Singapore; Tan Tock Seng Hospital, Singapore, Singapore; Lee Kong Chian School of Medicine, Nanyang Technological University, Singapore, Singapore

**Keywords:** SARS-CoV-2, COVID-19, Multi-arm Multi-stage, platform trials

## Abstract

**Background:** Safe and effective therapies for COVID-19 are urgently needed. In order to meet this need, the Accelerating COVID-19 Therapeutic Interventions and Vaccines (ACTIV) public-private partnership initiated the Therapeutics for Inpatients with COVID-19 (TICO). TICO is a multi-arm, multi-stage (MAMS) platform master protocol, which facilitates the rapid evaluation of the safety and efficacy of novel candidate anti-viral therapeutic agents for adults hospitalized with COVID-19. Four agents have so far entered the protocol, with rapid answers already provided for three of these. Other agents are expected to enter the protocol throughout 2021. This protocol contains a number of key design and implementation features that, along with challenges faced by the protocol team, are presented and discussed.

**Protocol Design and Implementation:** Three clinical trial networks, encompassing a global network of clinical sites, participated in the protocol development and implementation. TICO utilizes a MAMS design with an agile and robust approach to futility and safety evaluation at 300 patients enrolled, with subsequent expansion to full sample size and an expanded target population if the agent shows an acceptable safety profile and evidence of efficacy. Rapid recruitment to multiple agents is enabled through the sharing of placebo as well as the confining of agent-specific information to protocol appendices, and modular consent forms. In collaboration with the Food and Drug Administration, a thorough safety data collection and DSMB schedule was developed for the study of agents with limited in-human data.

**Challenges:** Challenges included ensuring drug supply and reliable recruitment allowing for changing infection rates across the global network of sites, the need to balance the collection of data and samples without overburdening clinical staff, and obtaining regulatory approvals across a global network of sites.

**Conclusion:** Through a robust multi-network partnership, the TICO protocol has been successfully used across a global network of sites for rapid generation of efficacy data on multiple novel antiviral agents. The protocol design and implementation features used in this protocol, and the approaches to address challenges, will have broader applicability. Mechanisms to facilitate improved communication and harmonization among country-specific regulatory bodies are required.

## 1 Background

There is an urgent need for novel and effective antivirals against SARS-CoV-2 to reduce the substantial morbidity and mortality seen with COVID-19. To address this need, the Accelerating COVID-19 Therapeutic Interventions and Vaccines (ACTIV) public-private partnership ^1^ selected three clinical trial networks, the International Network for Strategic Initiatives in Global HIV Trials (INSIGHT) ^2^, the Cardiothoracic Surgical Trials Network (CTSN) ^3^ and the Prevention and Early Treatment of Acute Lung Injury network (PETAL) ^4^ to collaborate, design and implement the ACTIV-3 protocol (Therapeutics for Inpatients with COVID-19 (TICO)). Given the urgent clinical need and the large number of emerging anti-SARS-CoV-2 therapeutic agents to be tested, the protocol team opted for a multi-arm multi-stage (MAMS) platform master protocol design. Efficiencies of the MAMS platforms include the ability to share/pool placebo controls across multiple agents, the use of intermediate efficacy futility and safety assessments such that only the most promising agents go forward into full enrollment, and the less promising are rejected early, thus avoiding overlapping or redundant work on parallel protocols, while maintaining scientific rigor including double blinding, randomization, placebo control, using a single database and regular reviews of interim data by an independent Data and Safety Monitoring Board (DSMB) and provide guidelines for early termination based on group sequential methods ^5, 6^. These features ensure the most efficient use of already stretched clinical, and regulatory resources.

While similar designs have been used successfully in many different settings, including during the current pandemic (e.g. RECOVERY trial (NCT04381936); WHO SOLIDARITY trial (ISRCTN83971151)), these studies have primarily studied re-purposed agents with relatively well-established safety profiles. TICO, however, was intended to provide rapid efficacy and safety data for novel antiviral agents in hospitalized patients, and to enable downstream drug regulatory approvals if an agent shows efficacy. Facilitated by a successful multi-network partnership and U.S Food and Drug Administration (FDA) collaboration, the protocol was designed and implemented rapidly (9 weeks from first protocol meeting to first participant randomised). So far, the TICO master protocol has been approved in eight countries and has generated results for three novel agents, LYCoV555 ^7^ (Eli Lilly and Company), Vir-7831 ^8^ (GlaxoSmithKline and Vir Biotechnology), and Brii-196/198 ^8^ (Brii Biosciences Limited) (Figure 1). Another agent, AZD7442 (AstraZeneca) remains under study, with further agents and countries poised to enter the protocol throughout 2021. There were a number of key design and implementation features of the TICO master protocol that enabled the rapid recruitment and results generated by this protocol. These features, along with challenges faced by the protocol team (Table 1), are presented and discussed here.

**Table 1.** Challenges in protocol design and implementation

| Challenges | Implemented or proposed solutions |
| --- | --- |
| Ensuring drug supply across a global network of sites | Request for waiving of relabelling requirements<br>Centralised drug management and distribution through two drug depots<br>Pragmatic registration and activation<br>Use of centralised pharmacies to serve multiple sites |
| Data and sample collection during a pandemic | Reducing reporting burden through the use of protocol specified exempt events<br>Attempting to balance safety reporting with burden on site staff<br>Phone visits for discharged patients<br>Use of contractors for post-discharge sample collection |
| Regulatory approval and study implementation outside the U.S. | Sharing of regulators responses across the network<br>Use of a master protocol design with all new agent specific changes in an appendix<br>Mechanism to allow communication between relevant agencies in different countries |

**Figure 1.**
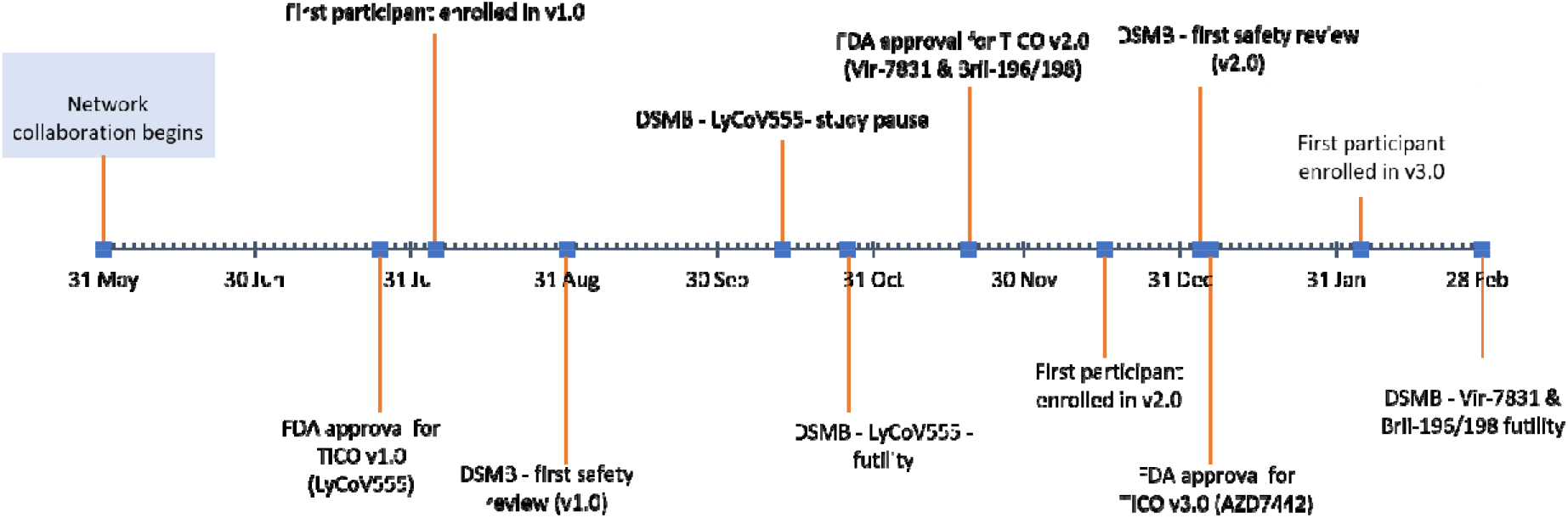
Project Timeline and Milestones. So far there have been three versions of the TICO protocol. Each version adds a new agent or agents. V1.0 included the Lilly neutralizing monoclonal anti-body LyCoV555. V2.0 of the protocol included the GSK/Vir neutralizing monoclonal antibody Vir-7831 and the Brii Bioscience neutralizing monoclonal antibodies Brii-196/198. V4 of the protocol included the AstraZeneca neutralising monoclonal antibody AZD7442. Key milestones for these protocol versions, including FDA approval, first participant enrolled, first safety review and futility, are presented in the figure.

## 2 Protocol Design and Implementation

### 2.1 Protocol oversight and Network Integration

The National Institute of Allergy and Infectious Diseases (NIAID) serves as the overall sponsor. Sites outside the U.S. are sponsored by the University of Minnesota. The Trial Oversight Committee (TOC) has been established to provide oversight for both the ACTIV-2 (NCT04518410) and ACTIV-3 initiatives and includes the trial co-chairs and representatives from Operation Warp Speed (OWS) therapeutics and NIAID. Additional voting members include leaders from National Heart, Lung and Blood institute (NHLBI), NIAID, Biomedical Advanced Research and Development Authority, FDA and the National Center for Advancing Translational Sciences (NCATS). The TOC also has responsibility for approving agents for entry into the TICO protocol, based on recommendations from the ACTIV agent selection committee (ASC). Candidate agents are submitted for consideration for TICO through a public portal, before undergoing a systematic scientific review by the ASC. The TOC votes on whether an agent enters TICO and considers a number of factors, including safety, *in vitro* potency against the virus, potential for viral resistance to arise, target epitope and potency (if the agent is an antibody), scale-up potential and dose and route of administration. ACTIV leadership requested TICO focus initially on neutralising monoclonal antibodies, with expansion to other novel antiviral agents as these become available. The TICO protocol team (see supplemental materials) is responsible for scientific and operational oversight. Implementation is coordinated by the INSIGHT Coordinating Centre (CC) at the University of Minnesota in collaboration with eight International Coordinating Centres (ICCs; five from INSIGHT and three representing the other networks). All have extensive experience managing clinical trials and work with >300 sites across North and South America, Europe, Australia, Africa and Asia. This large diverse network is important for three reasons. Firstly, a large global network is essential for recruitment, especially as case rates during the pandemic fluctuate regionally in unpredictable ways. Secondly, a broad range of clinical sites across multiple countries and continents results in a demographically diverse study population, ensuring any beneficial treatments identified have broad applicability. Thirdly, standard approaches for operations and trial conduct naturally vary across networks, and through collaboration the most effective and efficient from each network can be elevated and disseminated as ‘best practice’ across the full collaborative network.

In order to facilitate rapid approval and implementation of the protocol across the diverse network, certain roles and responsibilities were distributed to the ICCs, with central oversight by the CC. The CC managed drug distribution (in collaboration with PCI pharma services), central specimen storage and lab kit distribution (in collaboration with Advanced Biomedical Laboratories) and acted as the Statistical Data Management Centre. Each ICC is then responsible for the implementation and management of clinical research sites within their networks, including registration, regulatory approval, site training, lab kit ordering, drug orders, monitoring and ensuring data quality. To further facilitate implementation, ICCs often utilize in-country hubs, called Site Coordinating Centres (SCCs), who have extensive experience with regulatory and other requirements unique to their network of sites. See Supplemental Table 1 for details on the ICCs, SCCs and participating TICO sites.

### 2.2 Multi-arm, Multi-stage design of TICO

TICO is designed as a randomized, double blinded, placebo-controlled phase III Multi-arm Multi-stage (MAMS) platform master protocol. For any agent, at the outset of the trial, only participants without end-organ disease (Disease Stratum 1) will be enrolled. This more restricted enrollment will continue until approximately 150 participants per study arm are enrolled and followed for 5 days. At this point, the DSMB will carry out a pre-specified assessment of futility, based on two 7-category ordinal outcomes (pulmonary and pulmonary+), assessed at Day 5. Safety of the investigational agents will also be assessed. For investigational agents passing this initial futility assessment, enrolment will expand seamlessly, without any unblinding of data, to also include patients with end organ disease (Disease Stratum 2) The target population is narrower initially to expedite identification of early signals of safety and efficacy as patients with end organ dysfunction are unlikely to recover over 5 days and assessment of safety is more challenging. The expansion to include more severely ill participants is contingent on FDA and DSMB recommendations. If the initial futility assessment is passed, interim analyses are based on the primary endpoint of sustained recovery and use pre-specified guidelines to determine early evidence of benefit, harm or futility for the investigational agent.Once the full sample size is reached (estimated to be 1000 participants, equally allocated to each investigational agent and placebo), a confirmatory efficacy and safety analysis will take place (Figure 2). Procedures for data collection and primary endpoint ascertainment do not change for agents that pass the futility assessment, and all patients recruited prior to the futility assessment are included in the final efficacy assessment. For more details on the intermediate outcomes, primary endpoint and statistical analysis plan, see supplementary materials.

**Figure 2.**
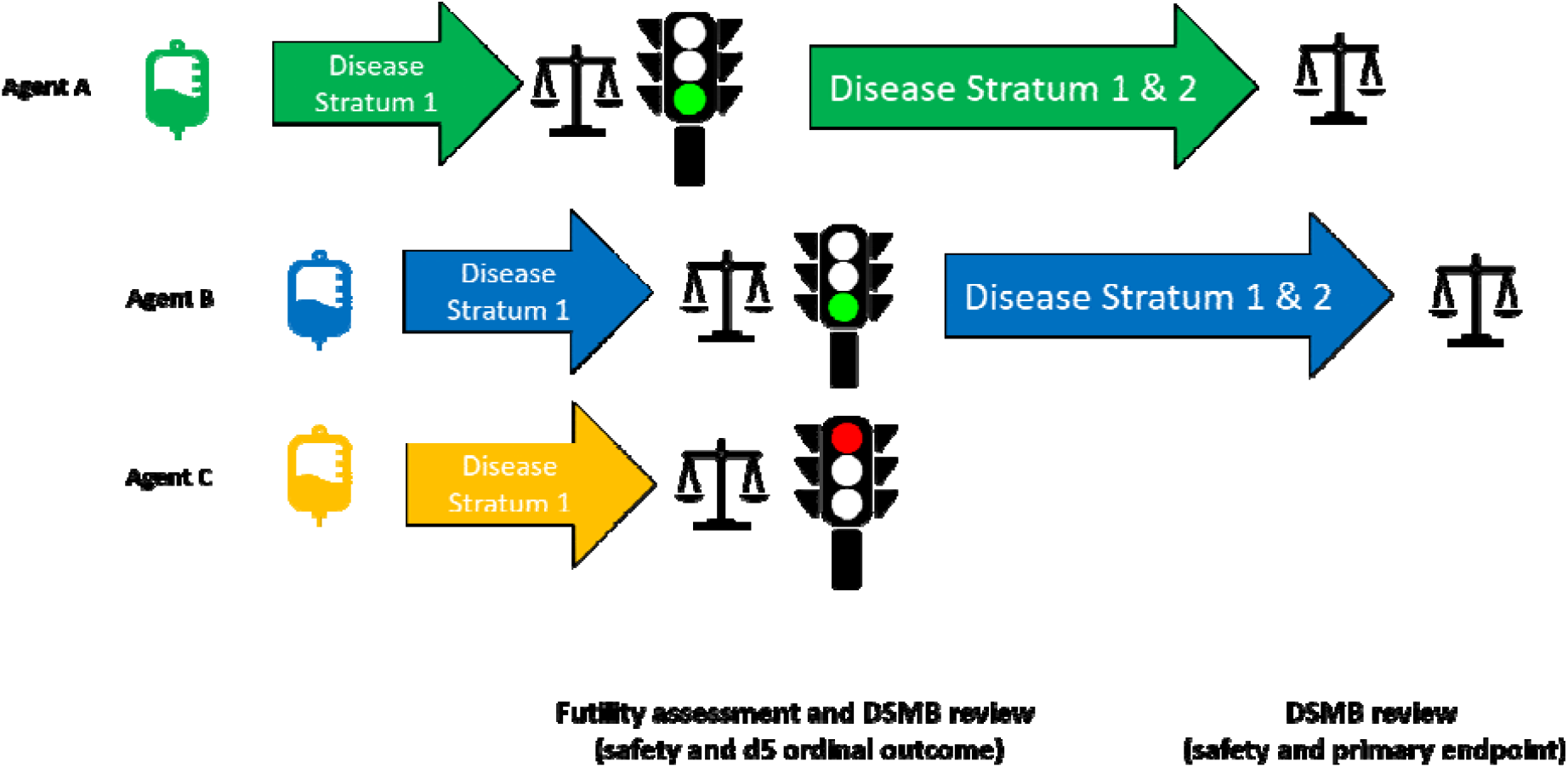
Agent entry and progression through the two-stages of the TICO study. The TICO study allows for multiple agents to be studied concurrently and for agents to enter the study at different time-points. In the theoretical scenario presented in figure 2, Agent A is the only agent that is available for randomisation at the beginning of the study. Later, Agent B and Agent C enter the study, and new participants are able to be randomized to all three agents (and placebo). Agent A completes recruitment in Disease Stratum 1 and, after DSMB review, is approved to also include those in Disease Stratum 2. Agent B and Agent C both complete recruitment in Disease Stratum 1, however, while Agent B proceeds, Agent C does not receive DSMB approval to proceed and randomisation to this agent ceases. Agent A and Agent B progress through Disease Stratum 2 and both undergo a final DSMB review of safety and efficacy (using the primary endpoint) when recruitment is complete.

### 2.3 Use of shared placebo in TICO

In TICO, each randomised participant could potentially receive any of the active agents for which they are eligible. The placebo group is then “pooled” so those randomized to the placebo of one agent will also serve as control for other agents to which the person could have been allocated. Thus, the probability of being allocated to any one investigational agent is the same as being allocated to placebo. The more concurrent agents under study at any given time therefore increases the probability of a participant being randomized to an active agent while also reduces the total participants required for an agent to reach crucial milestones. For example, the first agent to be studied using this protocol, LYCoV555 entered the protocol by itself and 314 participants were recruited over a 10-week period prior to the futility assessment. The second two agents, the Vir-7831 and Brii-196/198, began concurrently and were able to share placebo. At the time of the futility assessment for these agents, ∼11 weeks after first patient recruited, 168 participants had available day-5 data for Vir-7831, 166 for Brii-196/198 and 173 for placebo. If placebo were not shared, another 100 participants would have been needed for the futility assessment, costing time and resources, for the same result. However, it is worth noting that if by chance the shared placebo group was atypical e.g. having poorer prognosis than expected, the baseline imbalance would affect more than one trial, although this could be rectified by post hoc adjustment of the comparison between active and placebo in the affected trials.

### 2.4 Separate appendices for investigational agents and modular consent forms

Key to the success of TICO was the ability for multiple agents to be studied concurrently and for new agents to enter the protocol seamlessly. To facilitate this, the master protocol itself contained all relevant information and study procedures applicable to the broad conduct of the trial. All agent-specific information (including unique eligibility criteria, if any) are inserted into individual appendices. Thus, the entry of a new agent simply involves review of a new appendix by regulatory bodies and ethical boards, and the master protocol remains intact. This approach coupled with a modular information statement and consent form, with additional information sheets on individual drugs, and their side-effect profile, minimizes duplication for regulatory and site staff.

In instances where an individual cannot or should not be randomized to one or several of the agents (e.g. if agent specific eligibility criteria excludes them or an agent is unavailable due to pending regulatory approval, supply-chain or storage issues), we added two further key features. First, a randomization application was developed that factors in potential differences in both availability of study product and eligibility criteria between agents (see supplemental materials). Second the use of modular consents, as described above, easily allows investigators to inform participants which agents they may receive and then present the appropriate drug information.

### 2.5 Safety data collection and DSMB schedule for the study of novel agents

Many of the agents to be studied in TICO have limited in-human safety data. To ensure patient safety and adequate capture of data for future emergency use authorisation (EUA) and/or new drug application, the FDA reviewed and provided feedback on the protocol and DSMB schedule. As guided by the FDA, the specific safety collection (Supplemental Table 1) includes infusion-related reactions, targeted day 5 laboratory results (centrally graded) along with frequent assessments of AEs, serious adverse events (SAEs), and unexpected problems while hospitalised and post-discharge. For the first agent, participants were followed for 90 days. For next three agents, follow-up was extended to 18 months, due to longer half-lives for the new agents. The data collection beyond 90 days is restricted to death and hospitalizations, which was judged to provide sufficient information for safety monitoring without overburdening site staff. To review these safety data, and ensure safety of participants throughout the protocol, the DSMB conducts regular meetings while an agent is under study including a very early review (after 20-30 participants have day-5 data) and at the early futility assessment, and subsequent futility assessments (for more details see page 9 of the supplemental statistical analysis plan). The DSMB also receives weekly safety reports and can choose to convene additional meetings should concerning safety signals emerge.

## 3 Challenges in protocol design and implementation

### 3.1 Ensuring drug supply across a global network of sites

A major challenge faced was ensuring timely drug supply across a global network of sites to match the dynamic infection rates across geographical areas. A number of strategies were implemented to overcome this challenge. First, regulatory bodies were asked to waive the requirements to relabel study drugs, including translation into local languages, according to the local regulatory requirements. Secondly, drug distribution was centralized to two drug depots (one in the U.S. and one in the UK which later moved to Ireland), and the CC and ICCs closely monitored drug supply at individual sites through a central drug management database. This allowed the protocol team to monitor drug supply closely and send additional product to sites in need. Despite this, drug shipment to non-US and non-European study sites remain hampered by freight availability. Thirdly, in an attempt to best utilize the global network of sites and respond to the changing nature of global infection rates TICO registers all sites proactively, when all appropriate regulatory, registration and training documentation is in place, but only activates a site and ships study product when there is evidence or expectation of local disease activity. Finally, as infection rates and recruitment capabilities vary even across the same country/city, clinical sites are encouraged to select a pharmacy that can serve multiple clinical sites within a close geographical area, as opposed to a more traditional one-site one-pharmacy model (see Pharmacy Options in Supplemental materials). This one-pharmacy, multiple-sites model has resulted in efficient drug-distribution and reduced waste. Notable successes of this model were at CHIP, Rigshospitalet, Copenhagen (one-pharmacy, 10-sites), Duke University (one-pharmacy, three-sites), UCSF (one-pharmacy, two-sites) and Cleveland Clinic Foundation (one-pharmacy, three-sites).

### 3.2 Real-time data and sample collection during a pandemic

Detailed and well-standardized data collection during and after hospitalization (including sample collection and regular assessments of the primary endpoint) is essential for the regular safety and clinical efficacy assessments, as well as any future EUA or new drug applications. Due to local surges in case numbers during the pandemic, however, extensive data collection carried the danger of overwhelming the research staff at affected clinical sites, with health care worker infections exacerbating the situation. Further, stringent infection control measures posed challenges for patient review and sample collection, particularly post-discharge.

To reduce the burden on site staff, data collection was carefully calibrated. For example, clinical events that were already captured as part of the ordinal outcomes or other secondary objectives were exempt from additional SAE reporting (unless deemed related to an investigational agent). These “protocol specified exempt events” were defined in the protocol. Further, after day-7, AEs of any grade were collected as a snapshot on day 14 and day 28 only, while grade 3 and 4 AEs were collected retrospectively on day 14 and day 28). Longer term follow-up (after 90 days) was limited to vital status and hospitalisation only, which, as described above, was a balance between capturing key outcomes without overwhelming the clinical sites. Finally, some of the post-discharge study assessments were preformed over the phone, and contractors were hired to visit participants at home for post-discharge sample collection.

### 3.3 Regulatory approval and study implementation outside the U.S

Due to the involvement of the U.S. FDA, and a central ethics review by Advarra^®^, study implementation was rapid within the U.S.. However, regulatory approval and study implementation outside of the U.S. occurred slower and was a major challenge for the protocol team. For example, in the LYCoV555 substudy, only Denmark, Spain, U.K. and Singapore received approval by both ethics and medicines agencies by the time of the futility assessment, and only Denmark and Singapore opened in time to recruit.

There were three main reasons for these delays. Firstly, submission of the protocol to countries outside the U.S. required approval by both the FDA Advarra^®^ before the submission process could even begin. Secondly, due to the huge increase in COVID-19 related projects, many countries were facing a backlog of COVID-19 clinical trials applications and fast-track systems developed during the early phase of the pandemic became overwhelmed. Thirdly, regulatory agencies were reviewing data on novel antiviral agents for the first time and this necessitates careful review. Often, responses to these reviews required input from the pharmaceutical companies (specifically around pre-clinical data included in the agent’s submission data), which further delayed approvals.

A number of strategies were implemented to speed up regulatory reviews, including sharing of responses across ICCs to more swiftly deal with common questions and the use of SCCs to better coordinate submissions in specific countries. Future versions of the protocol may proceed more swiftly as regulatory agencies will only need to review the additional appendix with no major changes to the master protocol. However, global recruitment into large platform trials has the potential to substantially speed up the development of new treatments in a pandemic and ways to improve global implementation should be prioritized moving forward. One such improvement would be a formal mechanism that allows sharing of reviews between regulatory agencies (particularly between the FDA and other agencies). This way, the regulatory agency for each new participating country would have the benefit of communicating with other regulatory bodies and reviewing prior approvals and additional requested data. The intent would be to avoid repeated questions, give more confidence to the reviewing agency and generally speed up reviews.

## 4 Conclusion

The TICO master protocol responds to the urgent need to accelerate the development of safe, efficacious, novel antivirals for hospitalized COVID-19 patients. Through a successful collaboration of clinical trial networks, TICO has been rapidly and successfully designed and implemented globally. TICO is an efficient, flexible, rigorous MAMS platform master protocol that allows for concurrent safety and efficacy evaluation of multiple novel antiviral agents, with agents able to enter at different times. The use of an early futility assessment allows for the rapid selection of only the most promising agents for full evaluation using a clinically relevant primary endpoint, and therefore quickly removing agents from the trial that fail to demonstrate potential efficacy. Furthermore, the thorough safety data collection and frequent DSMB reviews allow speed and safety to co-exist. The study is currently underway in multiple countries and can respond to fluctuations in infection and recruitment rates across geographical areas. However, future trials would benefit from a process that would allow more rapid and efficient regulatory approval outside of the U.S. The unique design and implementation features of this protocol may inform future protocol design during the COVID-19 pandemic and in infectious diseases or acute respiratory failure research more broadly.

## Supporting information

Supplemental Statistical Analysis Plan

## Data Availability

There is no data relevant to this manuscript

## 5 Acknowledgements

The authors would like to thank the large number individuals across all the contributing trials networks, government agencies, local clinical sites, laboratory staff, pharmacists, logistics personnel, institutional review boards and regulatory bodies who contributed to the design and implementation of the TICO protocol.

TICO is funded primarily through Operation Warp Speed (now White House COVID-19 Response Team) sub-contracted through Leidos Biomedical Research Inc.. Additional funding was provided by the NIH (including NHLBI, NIAID), U.S. Department of Veterans Affairs, as well as the governments of Denmark (National Research Foundation; grant no 126), Australia (National Health and Medical Research Council), and U.K. (Medical Research Council, MRC_UU_12023/23).

