## Supplemental Materials for "Design and implementation of an international, multi-arm, multi-stage platform master protocol for trials of novel SARS-CoV-2 antiviral agents: Therapeutics for Inpatients with COVID-19 (TICO/ACTIV-3)"

### 1 Protocol team structure

To oversee the implementation of this master protocol, a protocol team was formed including: Protocol co-chair(s)

- NIAID, Division of Clinical Research representatives
- INSIGHT University of Minnesota representatives
- INSIGHT International Coordinating Center representatives
- Representatives from collaborating trials networks (i.e. PETAL, CTSN and the VA)
- Representative from ACTIV-2 protocol team
- Representatives from the central specimen repository
- Representative from the drug distribution group
- Representatives from collaborating manufacturers of investigational agents
- Representatives from site investigators
- Community representative(s)

A core team consisting of the co-chair(s), ICC leaders, NIAID representatives, study statisticians, representatives from collaborating trials networks, and other representatives and the INSIGHT PI will also regularly convene to review study progress and address study conduct and administrative issues that arise.

### 2 Primary objective and choice of a patient-centered primary endpoint

The TICO primary objective is to determine whether investigational agents are safe and efficacious compared with placebo when given with established standard of care (SOC). The primary efficacy endpoint is time to sustained recovery through day 90 i.e. when a participant is discharged from hospitalization to home and remains at home for at least 14 consecutive days. This patient-centered endpoint was chosen because of the extended duration of health impairment associated with COVID-19 ^[1-3]^. The longer follow-up to capture this endpoint (compared to the common 28 days ^[4-6]^) was designed to provide a more comprehensive assessment of the capacity of a therapeutic agent to speed recovery from COVID-19.

The TICO primary endpoint of sustained recovery is defined as 14 continuous days at home, where home is defined as the type or level of residence where the participant lived prior to their SARS-CoV-2 infection. This approach avoids categorizing patients as recovered if they continue to have care needs beyond their pre-morbid state despite discharge from an acute care facility, or if they are re-admitted to hospital shortly after initial discharge. To operationalize the collection of this endpoint, a participant’s ‘home’ is classified at enrolment (types of residences are defined below) and a participant’s current location, and consecutive days spent at that location, is collected fortnightly during follow-up using a dedicated CRF.

There are seven possible categories for classifying home in the TICO study. They are:

**Independent dwelling withOUT professional medical help -** Participant is living in a house, apartment, flat, condominium independently (regardless whether alone or with family or friends; also regardless of any paid help such as housekeeping service, maid, gardener etc.).

**Independent dwelling WITH professional medical help -** Participant is living in a house of any form, apartment, flat, or condominium but is requiring visiting professional medical help (e.g., visiting nurse, physiotherapist, or other home healthcare personnel meant to provide medical or rehabilitation care in the home)

**Community dwelling -** Participant is homeless, living on the streets or undomiciled, or may be living in a shelter or hotel (including hotel stay for quarantine purposes).

**Residential care facility -** These are non-skilled nursing facilities where care and services are provided to assist with activities of daily living. If the nature of the services can be safely and effectively performed by a trained nonmedical person, the services will be considered residential care. Examples include assisted living facility, group home, low-level care facility, or other nonmedical institutional setting.

**Other Healthcare facility -** Skilled nursing facility (nursing homes), acute inpatient rehabilitation facilities (acute rehab), or other healthcare facility that provides onsite medical care above a residential care facility but with a lower intensity than provided in hospitals.

**Long-term inpatient care hospital -** Long-term acute care hospital (LTACH), long-term care hospital. Note: These are hospitals/facilities meant to provide longer term (typically >20-30 days) of acute-care services after discharge from the short-term acute care hospital. Services requiring this level of care may include mechanical ventilation, intensive wound care, intensive pain management. LTACHs are hospitals that specialize in the treatment of patients with serious medical conditions that require care on an ongoing basis but no longer require intensive care or extensive diagnostic procedures.

**Short-term acute care hospital -** Short-term acute care hospital (similar to the index/enrolling hospital). Most acute care hospitals fall into this category, regardless of the duration of hospital admission.

### 3 Choice of intermediate outcomes for early futility assessment

Three key considerations drove the outcome selection for the futility and safety assessment: capacity to quickly assess for potential efficacy and safety, hypothesized high correlation with the primary endpoint of time to sustained recovery, and capacity to capture both pulmonary and non-pulmonary events among participants. Use of the primary endpoint for early futility and safety assessments was deemed impracticable, as it requires substantial follow-up time for ascertainment. Intermediate assessments must thus be made at much earlier time points, using surrogates for the ultimate primary endpoint. Given these design considerations, the protocol uses two ordinal outcomes at day 5 to determine whether an agent passes the futility assessment). The Pulmonary outcome is a 7-category outcome largely based on the degree of respiratory failure, adapted from a similar outcome used in the ACTT-1 study ^[5]^ and an initial WHO master protocol ^[7]^. A second ordinal outcome, called the Pulmonary-plus (Pulmonary+) outcome, adds extra-pulmonary conditions to the pulmonary outcome that cover a range of organ dysfunction associated with COVID-19. For each of the two outcomes, the highest category that applies on day 5 will be used. Agents that fail to meet a relatively modest bar for potential efficacy (p < 0.3 for one or both ordinal outcomes) or that exhibit concerning safety signals will not proceed to full efficacy assessment. More details can be found in the statistical analysis plan.

### 4  Randomization application

In order to facilitate randomizations to multiple possible agents, a flexible web-based randomization application was developed. The flexibility is accomplished with a database-driven approach pulling information from three tables: (i) randomisation table, which contains stratum specific schedules (as randomisation is stratified by pharmacy and disease severity) for one or multiple agents; (ii) drug table, which contains agent availability and allows stopping/restricting randomisation to selected agents, and information describing the agent, including number of doses of the agent available at the site study pharmacy; and (iii) constraint table, which contains contraindications and information used to modify inclusion/exclusion criteria. Randomisation assignments will be obtained in sequence from pre-generated schedules stratified by pharmacy and disease severity stratum. Allocation will be 1:1 Active:Placebo for one agent, 2:1:2:1 Active A:Placebo A:Active B:Placebo B for two agents (A and B), and so on. Using permuted blocks with k agents, every k placebo assignments will include one agent specific placebo assignment per agent, and every k active assignments will include one per agent. Using the mass-weighted urn scheme ^[8]^, the underlying Active:Placebo sequence is generated to ensure an approximate 1:1 balance for each active versus pooled placebo comparison within strata throughout the trial.

The application can also vary allocation according to stratum (i.e. pharmacy or disease severity). With 2 agents, allocation for the less severe stratum might be 2:1:2:1 as above but if agent B has not advanced to Disease Stratum 2 (and can therefore not recruit individuals with high disease severity), for the more severe stratum allocation would be 1:1 Active A: Placebo A. Furthermore, the application allows a limited number of sites to allocate patients 2:1:2:1: Active A:Placebo A:Active B:Placebo B or 1:1 Active B:Placebo B initially to obtain safety data for DSMB review for agent B while other sites randomize participants to only Active A; Placebo A until the safety review is complete.

### 5 Pharmacy set-up options

A number of pharmacy options are available to participating sites.

1. A single study site pharmacy serving multiple clinical sites within a close geographical area (e.g. the same city). Local site’s clinical staff screen and randomise patient before ordering relevant SOC and placebo/agent from the study site pharmacy. SOC and placebo/agent are made up and the placebo/agent is blinded at the study site pharmacy before being distributed to the local site clinical staff for administration.
2. A single study site pharmacy serving multiple local site pharmacies within a close geographical area. Local site’s clinical staff screen and randomise patient before ordering relevant SOC and placebo/agent from the study site pharmacy. The study site pharmacy selects the appropriate number of vials of both SOC and placebo/agent, but do not prepare the study product, to be transported to the LSP. At the LSP, the SOC and placebo are made up and the placebo/agent is blinded before being distributed to clinical staff for administration.
3. A traditional pharmacy set-up where the study site pharmacy only serves a single clinical site

### 6 Supplemental tables

Supplemental Table 1 Participating International Coordinating Centres (ICC), Clinical Sites and Site Coordinating Centres

| **INSIGHT Copenhagen ICC**  Centre of Excellence for Health, Immunity, and Infections (CHIP), Department of Infectious Diseases, Rigshospitalet, Copenhagen, Denmark | | |
| --- | --- | --- |
| **Site Name** | **City** | **Country** |
| University Hospital Zurich | Zurich | Switzerland |
| Unité VIH/SIDA Genèva | Geneva | Switzerland |
| Johann Wolfgang Goethe Univ. Ho sp., Infektionsambulanz CRS | Frankfurt | Germany |
| Universitätsklinik Köln | Cologne | Germany |
| Universitätsklinikum Regensburg | Regensburg | Germany |
| Hvidovre University Hospital, Department of Infectious Diseases | Hvidovre | Denmark |
| Aarhus Universitetshospital, Skejby | Aarhus | Denmark |
| Odense University Hospital | Odense | Denmark |
| Aalborg Hospital | Aalborg | Denmark |
| Rigshspitalet, Department of Infectious Diseases | Copenhagen | Denmark |
| Nordsjællands Hospital, Hillerød | Hillerod | Denmark |
| Zealand University Hospital Roskilde | Roskilde | Denmark |
| Kolding Sygehus | Kolding | Denmark |
| Herlev-Gentofte Hospital | Hellerup | Denmark |
| Bispebjerg Hospital | Copenhagen | Denmark |
| Wojewodzki Szpital Zakazny | Warsaw | Poland |
| Hospital Universitari Germans Trias i Pujol (site and INSIGHT Site Coordinating Centre Spain) | Badalona | Spain |
| Hospital General Universitario Gregorio Marañón | Madrid | Spain |
| Hospital Clínic de Barcelona | Barcelona | Spain |
| Hospital Universitario La Paz | Madrid | Spain |
| Hospital Clínico San Carlos | Madrid | Spain |
| Hospital del Mar | Barcelona | Spain |
| Hospital Universitari Vall d'Hebron | Barcelona | Spain |
| Hospital Universitario de Bellvitge | Hospitalet de Llobregat | Spain |
| AIDS and Clinical Immunology Research Center | Tbilisi | Georgia |
| Karolinska University Hospital | Stockholm | Sweden |
| **INSIGHT London ICC**  Medical Research Council Clinical Trials Unit at UCL, University College London, London, UK | | |
| **Site Name** | **City** | **Country** |
| Hôpital Saint-Louis | Paris | France |
| Groupe Hospitalier Sud Île de France | Melun | France |
| Hopital Lariboisière | Paris | France |
| Ospedale San Raffaele S.r.l. | Milan | Italy |
| L. Sacco Hospital-Institue of Infectious and Tropical Diseases | Milan | Italy |
| INMI Lazzaro Spallanzani IRCSS | Rome | Italy |
| Bergamo Hospital | Bergamo | Italy |
| Royal Sussex County Hospital | Brighton | United Kingdom |
| Royal Free Hospital | London | United Kingdom |
| Royal Victoria Infirmary | Newcastle upon Tyne | United Kingdom |
| Guy's & St. Thomas' NHS Foundation Trust | London | United Kingdom |
| Churchill Hospital | Oxford | United Kingdom |
| St James's University Hospital | Leeds | United Kingdom |
| Bradford Teaching Hospitals | Bradford | United Kingdom |
| MRC/UVRI Research Unit on AIDS (site and INSIGHT Site Coordinating Centre Uganda) | Entebbe | Uganda |
| Joint Clinical Research Center (JCRC) | Kampala | Uganda |
| Gulu Regional Referral Hospital | Gulu | Uganda |
| Mulago Hospital Complex | Kampala | Uganda |
| Lira Regional Referral Hospital | Lira | Uganda |
| Masaka Regional Referral Hospital | Masaka | Uganda |
| CISPOC | Maputo | Mozambique |
| National & Kapodistrian University of Athens Medical School (INSIGHT Site Coordinating Centre Greece) | Athens | Greece |
| Attikon University General Hospital | Athens | Greece |
| 1st Respiratory Medicine Dept, Athens University Medical School | Athens | Greece |
| AHEPA University Hospital | Thessaloniki | Greece |
| Dept of Critical Care and Pulmonary Medicine, Evangelismos General Hospital | Athens | Greece |
| Democritus University of Thrace | Alexandroupoli | Greece |
| 3rd Dept of Medicine, Medical School, NKUA | Athens | Greece |
| **INSIGHT Sydney ICC**  The Kirby Institute, University of New South Wales, Sydney, Australia | | |
| **Site Name** | **City** | **Country** |
| Hospital General de Agudos JM Ramos Mejia | Buenos Aires | Argentina |
| CEMIC | Buenos Aires | Argentina |
| Instituto Médico Platense | La Plata | Argentina |
| Fundación Arriarán | Santiago | Chile |
| St. Vincent's Hospital, Sydney | Sydney | Australia |
| Westmead Hospital | Sydney | Australia |
| NCGM | Tokyo | Japan |
| Fujita Health University Hospital | Toyoake | Japan |
| Tokyo Shinagawa Hospital | Tokyo | Japan |
| Tan Tock Seng Hospital | Singapore | Singapore |
| Chennai Antiviral Research and Treatment Clinical Research Site | Chennai | India |
| Institute of Human Virology-Nigeria (IHVN) | Abuja | Nigeria |
| Chulalongkorn University and The HIV-NAT | Pathumwan | Thailand |
| Bamrasnaradura Infections Diseases Institute | Nonthaburi | Thailand |
| Tel Aviv Sourasky Medical Center | Tel Aviv | Israel |
| Rambam Medical Center | Haifa | Israel |
| **INSIGHT Washington ICC**  Veterans Affairs Medical Center and George Washington University, Washington, DC, USA. | | |
| **Site Name** | **City** | **Country** |
| University of Illinois at Chicago | Chicago | United States |
| Washington DC VA Medical Center | Washington | United States |
| MedStar Health Research Institute | Washington | United States |
| Henry Ford Health System | Detroit | United States |
| Denver Public Health | Denver | United States |
| Cooper University Hospital | Camden | United States |
| Yale University School of Medicine | New Haven | United States |
| West Haven VA Medical Center | West Haven | United States |
| Hennepin Healthcare Research Institute/HCMC | Minneapolis | United States |
| NJMS Adult Clinical Research Center | Newark | United States |
| Hillsborough County Health Department, University of South Florida | Tampa | United States |
| SUNY Downstate Medical Center | Brooklyn | United States |
| Medical College of Wisconsin, Inc. | Milwaukee | United States |
| Lundquist Institute for Biomedical Innovation | Torrance | United States |
| Georgetown University | Washington | United States |
| UT Southwestern Medical Center | Dallas | United States |
| Parkland Health and Hospital Systems | Dallas | United States |
| Minneapolis VA Medical Center | Minneapolis | United States |
| Case Western Reserve University | Cleveland | United States |
| University of Minnesota | Minneapolis | United States |
| Instituto de Infectologia Emílio Ribas - IIER | Sao Paulo | Brazil |
| Center for Infectious Diseases at the UFES | Vitoria | Brazil |
| Complexo Hospitalar Professor Edgard Santos | Salvador | Brazil |
| Socios En Salud Sucursal Peru | Lima | Peru |
| Hospital Nacional Hipolito Unanue | El Agustino | Peru |
| Nutrición | Mexico City | Mexico |
| INER | Mexico City | Mexico |
| GEA | Mexico City | Mexico |
| Hospital General Dr. Aurelio Valdivieso | Oaxaca City | Mexico |
| **INSIGHT NIH-DCR ICC**  Department of Clinical Research, National Institute of Allergy and Infectious Diseases, Bethesda, MD, USA | | |
| **Country** | **Country** | **Country** |
| Lincoln Medical Center | Bronx | United States |
| Our Lady of the Lake Regional Medical Center | Baton Rouge | United States |
| Triple O Research Institute | West Palm Beach | United States |
| Maimonides Medical Center | Brooklyn | United States |
| CHRISTUS Spohn Shoreline Hospital | Corpus Christi | United States |
| Hendrick Medical Center | Abilene | United States |
| Baton Rouge General Medical Center | Baton Rouge | United States |
| Carilion Clinic | Roanoke | United States |
| Hoag Memorial Hospital Presbyterian | Newport Beach | United States |
| Cotton O'Neil Clinical Research Center | Topeka | United States |
| CHRISTUS Good Shepherd Medical Center | Longview | United States |
| Columbia University Medical Center | New York | United States |
| Velocity Chula Vista | Chula Vista | United States |
| Velocity San Diego | La Mesa | United States |
| Rhode Island Hospital | Providence | United States |
| Franciscan Health Indianapolis | Indianapolis | United States |
| Franciscan Health Michigan City | Michigan City | United States |
| The Miriam Hospital | Providence | United States |
| Memorial Healthcare System | Hollywood | United States |
| **INSIGHT U.S. Department of Veterans Affairs (VA) research network ICC** | | |
| **Site Name** | **Site City** | **Country** |
| West Los Angeles VA Medical Center | Los Angeles | United States |
| San Francisco VAMC | San Francisco | United States |
| Miami VAMC | Miami | United States |
| Bay Pines VAMC | Bay Pines | United States |
| Palo Alto VAMC | Palo Alto | United States |
| Houston VAMC | Houston | United States |
| Southern Arizona VA Healthcare System | Tucson | United States |
| Aurora VAMC | Aurora | United States |
| North Florida/South Georgia Veterans Health Sysem | Gainesville | United States |
| Edward Hines VA Hospital | Hines | United States |
| VA Southern Nevada Healthcare System | Las Vegas | United States |
| Salem VA Medical Center | Salem | United States |
| Ralph H. Johnson VA Medical Center | Charleston | United States |
| VA San Diego Healthcare System | San Diego | United States |
| VA Loma Linda Healthcare System | Loma Linda | United States |
| CJ Zablocki VA Med Center | Milwaukee | United States |
| Oklahoma City VA Health Care System | Oklahoma City | United States |
| VA Pittsburgh | Pittsburgh | United States |
| Richard L. Roudebush VA Medical Center | Indianapolis | United States |
| VA TVHS Nashville Campus | Nashville | United States |
| Sacramento VA Medical Center | Mather | United States |
| Portland VA Health Care System | Portland | United States |
| VA Providence Healthcare System | Providence | United States |
| VA Long Beach Healthcare System | Long Beach | United States |
| Saint Louis VAMC | Saint Louis | United States |
| **Prevention and Early Treatment of Acute Lung Injury (PETAL) ICC**  Massachusetts General Hospital, Boston, USA | | |
| **Site Name** | **Site City** | **Country** |
| Baystate Medical Center (site and ALIGNE Site Coordinating Center) | Springfield | United States |
| Maine Medical Center | Portland | United States |
| U. Florida, Health Science Center & Shands | Gainesville | United States |
| Beth Israel Deaconess Medical Center (site and Boston Site Coordinating Centre) | Boston | United States |
| Massachusetts General Hospital | Boston | United States |
| University of Mississippi Medical Center | Jackson | United States |
| UCSF San Francisco (site and California Site Coordinating Centre) | San Francisco | United States |
| Ronald Reagan UCLA Medical Center | Los Angeles | United States |
| Stanford University Hospital & Clinics | Stanford | United States |
| UC Davis | Davis | United States |
| UCSF Fresno | Fresno | United States |
| University of Texas Health Science Center | Houston | United States |
| UCSF Medical Center at Mount Zion | San Francisco | United States |
| University of Colorado Hospital (site and Colorado Site Coordinating Centre) | Aurora | United States |
| National Jewish Health \| St. Joseph Hospital | Denver | United States |
| University of Michigan Medical Center (site and Michigan Site Coordinating Centre) | Ann Arbor | United States |
| Montefiore Medical Center Moses Hospital (site and Montefiore-Sinai Site Coordinating Centre) | Bronx | United States |
| Montefiore North | New York | United States |
| Montefiore Weiler | New York | United States |
| Banner University Medical Center Tucson | Tucson | United States |
| Cleveland Clinic Foundation | Cleveland | United States |
| University of Cincinnati Medical Center (site and Ohio Site Coordinating Centre) | Cincinnati | United States |
| Cleveland Clinic Fairview Campus | Cleveland | United States |
| Cleveland Clinic Marymount Campus | Cleveland | United States |
| Harborview Medical Center | Seattle | United States |
| Cedars-Sinai Medical Center | Los Angeles | United States |
| Oregon Health and Science University (site and Pacific Northwest Site Coordinating Centre) | Portland | United States |
| Swedish Hospital Cherry Hill | Seattle | United States |
| Swedish Hospital First Hill | Seattle | United States |
| University of Washington Medical Center | Seattle | United States |
| UPMC Presbyterian | Pittsburgh | United States |
| UPMC Magee | Pittsburgh | United States |
| UPMC Mercy | Pittsburgh | United States |
| UPMC Shadyside | Pittsburgh | United States |
| Wake Forest Baptist Health (site and Southeast Site Coordinating Centre) | Winston-Salem | United States |
| Medical University of South Carolina | Charleston | United States |
| University of Kentucky | Lexington | United States |
| Virginia Commonwealth University Health System | Richmond | United States |
| Intermountain Medical Center (Site and Utah Site Coordinating Centre) | Murray | United States |
| University of Utah Hospital | Salt Lake City | United States |
| Utah Valley Regional Medical Center | Provo | United States |
| LDS Hospital | Salt Lake City | United States |
| Vanderbilt University Medical Center | Nashville | United States |
| University Medical Center | New Orleans | United States |
| **Cardiothoracic Surgical Trials Network (CTSN) ICC**  Icahn School of Medicine at Mount Sinai, New York, USA | | |
| **Site Name** | **City** | **Country** |
| Allegheny General Hospital | Pittsburgh | United States |
| Baylor College of Medicine | Houston | United States |
| Baylor, Scott and White Health | Dallas | United States |
| Cedars-Sinai Medical Center | Los Angeles | United States |
| CHI St. Vincent, Arkansas | Little Rock | United States |
| Duke University Hospital | Durham | United States |
| East Carolina Heart Institute | Greenville | United States |
| Emory University | Atlanta | United States |
| Inova Heart & Vascular Institute | Falls Church | United States |
| Lutheran Medical Group | Fort Wayne | United States |
| MH Mission Hospital | Asheville | United States |
| Mount Sinai Medical Center | New York | United States |
| New York University Langone Health | New York | United States |
| Northwell Health | Manhasset | United States |
| Ochsner Clinic | New Orleans | United States |
| Piedmont Healthcare | Atlanta | United States |
| Texas Heart Institute | Houston | United States |
| University of Louisville | Louisville | United States |
| University of Maryland | Baltimore | United States |
| University of Southern California | Los Angeles | United States |
| University of Virginia Health Systems | Charlottesville | United States |
| WakeMed Heart Center | Raleigh | United States |
| West Virginia University | Morgantown | United States |
| Dartmouth-Hitchcock Medical Center | Lebanon | United States |
| Hôpital Laval | Quebec | Canada |

Supplemental Table 2 Safety Data Collection Schedule

|  | Infusion +2 hrs | Days 0-7 | Day 14 | Day 28 | Day 90 | Month 6, 12 and 18 |
| --- | --- | --- | --- | --- | --- | --- |
| Infusion-related reactions and symptoms | X |  |  |  |  |  |
| Incident grade 3 and 4 clinical AEs |  |  | X^1^ | X^1^ |  |  |
| Clinical AEs of any grade severity | X | X | X^2^ | X^2^ |  |  |
| Targeted laboratory abnormalities of any grade |  | X  (Day 5) |  |  |  |  |
| Hospital admissions and deaths | Collected through to Month 18 | | | | | |
| Serious AEs  (including those reported as part of the pulmonary and pulmonary+ ordinal outcomes) | Collected through Day 90 | | | | |  |
| Unanticipated problems | Collected through Day 90 | | | | |  |
| Any serious adverse event related to study intervention | Collected through Day 90 | | | | |  |

1. All grade 3 and 4 events since previous visit
2. All grade 1 and 2 events on the day of the visit only

1. Mitrani RD, Dabas N, Goldberger JJ. **COVID-19 cardiac injury: Implications for long-term surveillance and outcomes in survivors**. *Heart rhythm* 2020.

7. Organization WH. **COVID-19 Therapeutic Trial Synopsis**. In; 2020.

8. Zhao W. **Mass weighted urn design--A new randomization algorithm for unequal allocations**. *Contemp Clin Trials* 2015; 43:209-216.
