## Supplemental Statistical Analysis Plan for "Design and implementation of an international, multi-arm, multi-stage platform master protocol for trials of novel SARS-CoV-2 antiviral agents: Therapeutics for Inpatients with COVID-19 (TICO/ACTIV-3)"

**Version 2.0**

**A Multicenter, Adaptive, Randomized, Blinded Controlled Trial of the Safety and Efficacy of Investigational Therapeutics**

**for Hospitalized Patients with COVID-19**

Clinicaltrials.gov identifier: NCT04501978

EudraCT number: 2020-003278-37

| **Version** | **Date** | **Who** | **Comments** |
| --- | --- | --- | --- |
| 1.0 | 06 October 2020 | BG | TICO ACTIV-3 INSIGHT 014 Protocol v1.0, 27 July 2020  Investigational agent: LY-Cov555 |
| 2.0 | 28 January 2021 | BG | TICO ACTIV-3 INSIGHT 014 Protocol v2.0, 3 November 2020 and v3.0, 7 January 2021  Investigational agents: Vir-7831; BRII-196 and BRII-198; AZD7442 |

### 1 Introduction

#### 1.1 Objective of the Statistical Analysis Plan

The objective of this statistical analysis plan (SAP) is to provide a description of the general analytic strategy and the statistical methods that will be used to analyze the data for the TICO (Therapeutics for Inpatients with COVID-19) Phase III randomized, blinded, controlled, platform trial. This SAP applies to versions 2 and 3 of the TICO protocol. In version 2, two investigational agents are being studied, a SARS-CoV-2 neutralizing monoclonal antibody (nMAb) (Vir-7831) which is being developed by Vir Biotechnology (Vir) (San Francisco, CA) and GlaxoSmithKline (GSK) (Brentford, U.K.), and two nMAbs given sequentially (BRII-196 and BRII-198) which are being developed by Brii Biosciences (Durham, NC and Beijing). In protocol version 3, a third investigational agent is added, AZD7442, which is a combination of two nMAbs by AstraZeneca (Cambridge, U.K.). Participants are followed for death or re-hospitalization up to month 12 in version 2, and up to month 18 in version 3. Otherwise, the protocol versions are identical. The nMAbs are given by single infusion, or two sequential infusions for BRII-196 and BRII-198.

The primary objective of the platform trial is to determine whether investigational agents that are aimed at enhancing the host immune response to SARS-CoV-2 infection are safe and superior to control (e.g., placebo) when given with standard of care (SOC) for the primary endpoint of time to sustained recovery evaluated up to 90 days of follow-up.

In the platform trial, several agents may be investigated in parallel, or staggered with overlapping times; investigational agents may be added or dropped. When more than one agent is being tested concurrently, participants will be randomly allocated across agents (as well as between the agent and its matched placebo), and the control group is pooled across the concurrently randomized, agent-specific matched placebo groups. Thus, each investigational agent and the corresponding pooled control group form their own randomized trial, and several agents may (at least partially) share their pooled control groups.

This SAP:

- Provides a short description of the study design (sections 1.2-1.4)
- Describes goals of the interim reviews by the independent DSMB and the planned format of the review meetings (section 2)
- Describes the planned data analyses presented in the reports to the DSMB (sections 3-13). General analysis principles are summarized in section 3, safety analyses are described in section 7, efficacy analyses in section 8, and interim monitoring guidelines in section 9.
- Describes data summaries to be provided regularly to study leadership to aid in monitoring trial conduct and data quality; these data summaries will be pooled across treatment groups, and will be restricted to enrolment, baseline data, and summaries of data completeness and study conduct.

As needed, the overall SAP for TICO will be updated by the blinded study statisticians; it is planned to update the SAP in parallel with protocol amendments.

#### 1.2 Description of the Study Design

This section is adapted from Section 1 of the TICO protocol versions 2.0 and 3.0.

**Design**

TICO is a master protocol to evaluate the safety and efficacy of multiple investigational agents aimed at modifying the host immune response to severe acute respiratory syndrome coronavirus 2 (SARS-CoV-2) infection, or directly enhancing viral control in order to limit disease progression.

Trials within this protocol will be adaptive, randomized, blinded and initially placebo-controlled. Participants will receive standard of care (SOC) treatment as part of this protocol. If an investigational agent shows superiority over placebo, SOC for the study of future investigational agents may be modified accordingly.

The protocol is for a phase III randomized, blinded, controlled platform trial that allows investigational agents to be added and dropped during the course of the study for efficient testing of new agents against control (i.e., placebo + SOC) within the same trial infrastructure. When more than one agent is being tested concurrently, participants will be randomly allocated across agents (as well as between the agent and its placebo). For analysis, placebo groups of concurrently randomized agents will be pooled; therefore, control groups may overlap for different agents.

Randomization will be stratified by study site pharmacy and disease severity. There are 2 disease severity strata, defined as below:

- **Disease severity stratum 1:** Absence of all of the following: stroke, meningitis encephalitis, myelitis, myocardial infarction, myocarditis, pericarditis, symptomatic congestive heart failure (NYHA class III or IV), arterial or deep venous thrombosis or pulmonary embolism, requirement for invasive mechanical ventilation, ECMO, mechanical circulatory support, vasopressor therapy, or new renal replacement therapy.
  - Per FDA recommendation, none of the three investigational agents in version 3 of the protocol can be used in patients on high-flow oxygen or non-invasive ventilation. Therefore, all reference to “disease severity stratum 1” in this SAP also excludes such patients.
  - Initially, only patients in disease severity stratum 1 are eligible for enrolment, as described below.
- **Disease severity stratum 2:** Presence of at least one of the excluded conditions or treatments in disease severity stratum 1.

The **primary endpoint** is the time from randomization to sustained recovery, defined as being discharged from the index hospitalization, followed by being alive and home for 14 consecutive days prior to Day 90. The definition of home will be operationalized as the level of residence or facility where the participant was residing prior to hospital admission leading to enrollment in this protocol.

An independent Data and Safety Monitoring Board (DSMB) will regularly review interim analyses that summarize safety and efficacy outcomes. For any agent, at the outset of the trial, only participants in disease severity stratum 1 will be enrolled. This more restricted enrollment will continue until approximately 150 participants per study arm are enrolled and followed for 5 days. At this point, the DSMB will carry out a pre-specified assessment of futility, based on two 7-category ordinal outcomes (pulmonary and pulmonary+), assessed at Day 5. The pulmonary and pulmonary+ outcomes are described in Appendix A. Safety of the investigational agents will also be assessed.

For investigational agents passing this initial futility assessment, enrolment of patients in disease severity stratum 1 will continue, and it is planned to also expand enrollment, seamlessly and without any data unblinding, to include participants in disease severity stratum 2. The expansion to include more severely ill participants will be subject to recommendations by the FDA and the DSMB based on safety considerations.

After the initial futility assessment is passed, future interim analyses will be based on the primary endpoint of sustained recovery and will use pre-specified guidelines to determine early evidence of benefit, harm or futility for the investigational agent.

**Primary Objective**

The primary objective of this protocol is to determine whether investigational agents, initially focusing on those that are aimed at enhancing the host immune response to SARS-CoV-2 infection are safe and superior to control (e.g., placebo) when given with SOC for the primary endpoint of time to sustained recovery evaluated up to 90 days after randomization.

**Duration**

Participants will be followed for 18 months following randomization in version 3 of the protocol, 12 months in version 2. Primary and most secondary outcomes will be collected during the first 90 days of follow-up only. Follow-up beyond 90 days is planned because the half-lives of some agents indicate that potentially meaningful amounts may remain in the body after 90 days of follow-up. After 90 days through the end of follow-up, hospitalizations and deaths will be ascertained.

**Sample size**

This phase III trial is planned to provide 90% power to detect a 25% increase in the rate of sustained recovery for an investigational agent compared to placebo at the 0.025, 1-sided level of significance. This requires 843 primary events (i.e., participants who achieve sustained recovery). Randomization of 1,000 participants, equally allocated to each investigational agent and placebo, followed for 90 days is estimated to result in the required number of primary events. The event target may be achieved earlier if more than 1,000 participants are enrolled. Sample size will be evaluated periodically by study team members who are blinded to interim results on treatment difference and may be increased to maintain power for the hypothesized difference in sustained recovery between the investigational agent and placebo.

**Population**

The study population consists of inpatient adults (≥18 years) who have had COVID-19 symptoms ≤ 12 days. Initially, enrollment is restricted to disease severity stratum 1. After a pre-specified review by the DSMB for safety and futility, when approximately 150 participants are enrolled per study arm, eligibility for randomization will be expanded to also include patients in disease severity stratum 2, subject to recommendations by the FDA and DSMB.

**Stratification**

Randomization is stratified by study site pharmacy; once enrolment is expanded to also include disease severity stratum 2, randomization will also be stratified by disease severity stratum.

**Monitoring**

An independent DSMB will review interim data on a regular basis for safety and efficacy. An initial futility assessment will be performed after the first 300 participants (150 in the active and 150 in the placebo arms) are enrolled and have Day 5 data; this initial assessment is based on two ordinal outcomes (pulmonary and pulmonary+ outcomes at Day 5). Afterwards, the DSMB will use pre-specified guidelines to identify agents with clear evidence of efficacy for the primary outcome and, if so, recommend unblinding of the trial results for that agent. Conversely, the DSMB may recommend discontinuation of an investigational agent if the risks are judged to outweigh the benefits or if futility assessments indicate that there is low probability that an investigational agent will achieve statistical significance for the primary endpoint of sustained recovery.

For an investigational agent, if the trial is stopped early or if the trial continues until the pre-specified number of primary endpoints is reached, further enrollment of the investigational agent will be terminated if applicable, and the trial data for the investigational agent will be unblinded and reported with data through 90 days of follow-up. Follow-up of all participants will continue through 18 months (12 months in version 2.0 of the protocol) using the data collection plan described in the master protocol.

#### 1.3 Randomization

The randomization is described in section 6.1 of the protocol.

Patients will be equally allocated to each investigational agent + SOC or to placebo + SOC. For example, for a study of a single investigational agent, participants will be randomized in a 1:1 ratio to the investigational agent or placebo. If a patient is eligible for two investigational agents, the allocation will be 1:1:1 to investigational agent A, agent B, or placebo. Because the two investigational agents (A and B) may require different placebos (for example, when infusion volumes differ), the 1:1:1 allocation ratio will be achieved through a two-step randomization procedure: in *step 1*, the participant is randomized 2:1 to “active” versus “placebo”; in *step 2*, the participant is randomized 1:1 to A versus B. With k agents, this can be viewed as an initial k:1 allocation to “active” versus “placebo”, followed by a second, even allocation to one of the available agents (for example, if a participant was allocated to “placebo” in step 1, then the step 2 allocation will be 1:1 to “matched placebo for A” versus “matched placebo for B”). For the analysis, the concurrent agent-specific placebo groups will be pooled, resulting in a 1:1 allocation ratio for comparing each investigational agent versus the (pooled placebo) control group. If investigational agents are added or dropped, the allocation ratio to active versus placebo will be appropriately modified, and sample size will be recalculated as appropriate.

Randomization will be stratified by study site pharmacy (several clinical sites may share one pharmacy) and severity of disease at entry; the two disease severity strata are defined in section 1.2 of this SAP.

If more than one investigational agent is being compared with placebo and they have different contraindications, it is possible that a participant is eligible only for a subset of agents.

Comparisons will be of each investigational treatment against its control arm. The control arm consists of all participants who were “at risk” of being randomized to the investigational agent but were randomized to a control group instead. This concept is relevant when the randomization includes investigational agents with different eligibility criteria, when agents are introduced into the platform trial at different time points, or randomization to one of the agents is halted temporarily. Formal randomization includes agent-specific matched placebo groups, and the placebo groups will be pooled across agents, but only participants who 1) were eligible for the investigational agent under consideration, and 2) were randomized contemporaneously and at participating sites will be included in the control group for a given agent. At the time of randomization, for each participant, indicator variables will be set that record whether an agent was included in the randomization for that participant (e.g., indicator A=1, indicator B=1, indicator C=0 if the participant was eligible to be randomized to agents A and B, but not C). The pooled control group for agent A then consists of all participants who were randomized to (any) placebo, and for whom indicator A=1.

#### 1.4 Sample Size Estimates

The planned sample size for each pairwise comparison is 1,000 participants (500 participants in each group). The sample size is sufficient to detect a recovery rate ratio (RRR) of 1.25 for time to sustained recovery with 90% power, using a one-sided test with a significance level of 0.025. The treatment groups are compared using Gray’s test with ρ=0, the competing risks analogue of the log-rank test.

Sample size calculations are described in detail in Section 6.3 of the protocol.

Blinded sample size re-estimation will be carried out before enrollment is complete to determine whether the planned sample size of 1,000 participants followed for 90 days will yield the planned number of 843 primary events. The blinded sample size re-estimation does not involve unblinding of the treatment difference. It will be based on the pooled outcome data. Sample size may be increased to achieve the event target in the planned follow-up of 90 days. A sample size increase may also be considered in order to achieve the event target before all participants are followed for 90 days.

When 300 participants (150 per study arm) are enrolled and have Day 5 outcome data, an early futility assessment is planned. This assessment will be based on two ordinal outcomes, denoted as “pulmonary” and the “pulmonary+”; both are ordered categorical outcomes with 7 categories, assessed on Day 5 (the outcomes are described in Appendix A). Treatment groups will be compared using proportional odds models. With 300 participants, the early futility assessment is powered to detect a summary OR=1.60 with power of 95%, using a one-sided test with significance level of 0.30.^1^ Given the two-outcome decision rules, an investigational agent with a summary OR=1.60 at Day 5 for both outcomes, would pass the futility assessment with a power between 93% and 98%, and a type I error between 0.21 and 0.39.

### 2 Interim DSMB Reviews: Goals and Format

**Each investigational agent versus control will be reviewed as a separate clinical trial**; separate data reports will be prepared for each investigational agent and the corresponding randomized (pooled) placebo group.

**Goals of the interim reviews:**

- Protect the	safety of study participants.
- Advise on stopping or modifying the trial for efficacy, for patient safety in case of emerging data on harm, or for futility.
- After the first 300 participants are enrolled (150 participants per arm; all in disease severity stratum 1), advise on continuing the trial, and on expanding the study population to include more severely ill patients (disease severity stratum 2).
- Review the conduct of the trial
- If an investigational agent is stopped (due to efficacy, safety, or futility), the DSMB may be asked to advise on the timing of unblinding the data, in case the unblinding of the shared pooled placebo group may impact the integrity of the ongoing trial for another agent (section 14).

The DSMB will conduct frequent safety reviews. For an investigational agent with minimal pre-existing data, the first safety review will be conducted after 20-30 participants have been enrolled (10-15 per study arm) and Day 5 data are available, before increasing the pace of enrollment. Subsequent reviews will be timed according to the recommendations of the DSMB and study leadership. After the early futility review when 300 participants are enrolled (150 per study arm), further futility reviews would be expected to occur at approximately 50% and 75% information time (the number of observed sustained recoveries as a proportion of the targeted number of 843 events).

**The DSMB may request interim reports that are focused on safety at any time**.

**Review meetings** for each agent will typically consist of an Executive session (optional; closed), open session, closed session, and a second open session to give feedback to study leadership (optional). If several agents are reviewed at the same meeting, agents will be reviewed consecutively, either with a sequence of open and closed sessions, or with one open session and one closed session (provided there are no unblinding conflicts).

**Masking of treatment group labels in interim reports:** In the open reports, any data reports will be pooled across the two treatment groups (the specific investigational agent and its pooled control group as described above). In the closed reports, treatment group labels will masked; for example as “Group A” versus “Group B”. The treatment group labels will be consistent across all analyses and over subsequent reports. The DSMB will be unmasked to the treatment group labels.

**Open report to the DSMB**

The open reports for each investigational agent versus placebo comparison will contain:

- A synopsis of the trial design and current status of the platform trial
- Responses of the study team to DSMB requests
- A summary prepared by the study leadership
- Data summaries for enrolment, eligibility violations and protocol deviations, baseline characteristics
- Summary reports for data completeness and study conduct, pooled across treatment groups.
- Emerging external data, e.g., results of phase I or II trials on the investigational agent, will also be provided to the DSMB by the study leadership. This is usually included with the open report, but may be shared confidentially if needed.

All data summaries in the open report will be pooled across the investigational agent and placebo control. The open reports will be prepared by the blinded statisticians in cooperation with the unblinded statisticians. In addition to the DSMB, open reports will be provided to the study team, and posted on the website for access by study investigators.

While the study is ongoing, summaries by treatment group, and comparisons of the investigational agent versus placebo are restricted to the confidential closed report to the DSMB. Additionally, all summaries of follow-up data other than the data completeness and study conduct reports (pooled across the two treatment groups) will be restricted to the confidential closed report. For the **planned sample size re-estimations prior to completion**, the pooled number of primary events and the pooled event rate will be provided to the blinded study statisticians and study leadership. On a case-by-case basis, other pooled follow-up data may be provided if explicitly approved by the DSMB.

**Closed report to the DSMB**

All data summaries in the closed report will be by (masked) treatment group. The closed reports for a full review will contain:

- Specific data summaries requested by the DSMB or study leadership
- Data summaries in the open report, by treatment group (enrollment, baseline characteristics, eligibility violations)
- Data summaries to assess safety of the investigational treatment, described in sections 6 and 7. Data summaries for the primary “efficacy outcomes” and selected secondary outcomes will also be included in each report, because these data contain information about the risk/benefit profile of the investigational agent. Analyses are described in section 8.
- Data summaries on data completeness and study conduct, described in section 10
- Interim monitoring boundaries for efficacy or harm (section 9)
- Futility analyses (sections 9.2 and 9.4)
- Listings of grade 3 and 4 adverse events, serious adverse events (SAE), clinical organ failure and serious infections (PSEE), unanticipated problems (UP), suspected unexpected serious adverse reactions (SUSAR), and deaths.

**Data reports will follow a similar format for all investigational agents.** Each agent will have a small assigned team of unblinded statisticians, with 2-3 alternating teams when 2 or more agents are investigated in parallel. The unblinded statistician teams will cooperate in designing the master layout for the data reports and will serve as each other’s backup when needed. The unblinded statisticians will be unblinded to several investigational agents in the platform trial; those for which they serve as primary statisticians, and those for which they serve as backup or advisory statisticians.

### 3 Analysis Principles

**Each investigational agent versus control will be treated as a separate clinical trial**; data reports will be for one “target” investigational agent and its corresponding randomized (pooled) control group. Investigational agents will not be directly compared against each other, unless explicitly stated in the agent-specific data analysis plan and agreed upon by all stakeholders. Therefore, in the event that several investigational agents are included in the platform trial in parallel, the pairwise comparisons of each agent versus control will **not** be adjusted for potential inflation of Type I error “due to multiple comparisons”.

Comparisons will be of each investigational treatment against its (pooled) control arm.

**Analysis populations:**

- Comparisons for safety outcomes will be by modified intention-to-treat. The modified intention-to-treat analysis is restricted to participants who received a complete or partial infusion of the investigational agent/placebo; participants who did not receive any of the investigational agent/placebo are excluded.
- Comparisons for efficacy endpoints will be by intention-to-treat, unless otherwise stated. Sensitivity analyses by modified intention-to-treat will be carried out for primary outcomes and key secondary outcomes.

**Pooled control group:** As stated in section 1.3 above, the control arm for any investigational agent will be pooled across the agent-specific control groups for all agents that concurrently participated in the randomization. Specifically, the pooled control group for investigational agent A consists of all participants who might have been randomized to agent A but were randomized to a placebo group instead. This concept is relevant when a participant is eligible to be randomized to more than one investigational agent, and agents were introduced into the platform trial at different time points or have different eligibility criteria.

In order to identify the pooled control group for each investigational agent correctly, the randomization application is setting indicator variables at the time of randomization for each participant that record whether an agent was included in the randomization (e.g., indicator A=1, indicator B=1, indicator C=0 if the participant was eligible to be randomized to agents A and B, but not C). The pooled control group for agent A then consists of all participants who were randomized to (any) placebo, and for whom indicator A=1.

Therefore, only participants who 1) were eligible for the investigational agent under consideration, 2) were randomized contemporaneously and at participating sites, and 3) were randomized to placebo will be included in the control group for a given agent.

**Descriptive statistics** will be reported overall and by randomized group. For categorical outcomes, the number and percent in each category will be reported; percentages will be of non-missing values, if data are not complete. Continuous variables will be summarized by median (interquartile range [IQR]) and/or mean (SD). Continuous variables may be categorized (e.g., age may be broken into categories to investigate the distribution across age groups).

**Stratification:** Tests comparing the investigational agent versus control for primary outcomes and key secondary outcomes will be stratified according to the planned randomization strata (disease severity and site pharmacy), provided participant numbers are sufficiently large. In this analysis plan, “stratification by disease severity” refers to the two disease severity randomization strata described in section 1.2. Initially, participants are enrolled only in disease severity stratum 1, until the investigational agent has passed the initial futility assessment (when 300 participants are enrolled for the pairwise comparison; 150 per study arm) and has been approved to expand enrolment to include participants from both strata 1 and 2. In this SAP, we use the notation “stratification by disease severity and site pharmacy” to denote the following:

- For analyses that include only participants in disease severity stratum 1, analyses will be stratified by site pharmacy.
- For analyses that include participants in both disease severity strata, analyses will be stratified by site pharmacy within each disease severity stratum; this means, the maximal possible number of strata is twice the number of site pharmacies. For analyses where stratification is implemented through addition of indicator variables for strata to the model, the strata would be defined through main effects for disease severity, for site pharmacy, and the interaction between disease severity and site pharmacy.

Because there are many site pharmacies, some strata may be small, particularly early in the trial. In order to avoid loss of power, any stratum that contains too few participants (less than 10-20 participants or events) should be pooled with other strata (of the same disease severity, and preferably within the same country or geographical region). Thus, several small strata may be pooled together, or pooled with a larger stratum.

For time-to-event analyses, if strata are too small for fitting separate baseline hazard functions, strata may be added as a categorical covariate to models instead. Whenever possible, however, analyses should be stratified by disease severity (with separate baseline hazard functions).

For **binary outcomes**, probabilities will be compared between the investigational agent and its control group using Cochran-Mantel-Haenszel tests (CMH) or logistic regression. If the numbers are sufficiently large, CMH tests will be stratified according to the planned randomization strata (disease severity and site pharmacy), as described above under “stratification”. Odds ratios (OR) with 2-sided 95% confidence intervals (CI) will be estimated using logistic regression models.

For longitudinally measured binary outcomes, the treatment effect through follow-up will be estimated with 95% confidence intervals using generalized estimating equations (GEE) with a logit link function; the treatment effect is estimated via the interaction between the indicator for treatment group and the indicator for follow-up (versus baseline) visits. When there is more than one follow-up visit, “visit number” (day) may be included as categorical variable in the model, for variance reduction; alternatively, “time” may be included as a continuous variable.

**Ordered categorical outcomes** **(pulmonary and pulmonary+)** will be compared between treatment groups using proportional odds models, and the summary OR will be estimated with a 2-sided 95% CI.^2^ Additionally, to aid the interpretation, the ordinal outcome will be dichotomized according to cumulative probabilities of the ordered categories, comparing treatment groups for proportions of participants in category 1, in the “best 2 categories”, “best 3 categories”, etc.; these comparisons will be performed using logistic regression (or stratified CMH tests).

Models will be adjusted for the baseline categories of the pulmonary+ outcome and for study pharmacy, by including the corresponding indicator variables in the model.

- If the number of observations is too small to adjust for both categorical covariates, preference will be given to the adjustment for the pulmonary+ category at baseline. Site pharmacy categories may be collapsed as described above under “stratification”.
- For the initial futility analysis (after 150 participants per arm have Day 5 data), the adjustment for the pulmonary+ categories and pharmacy will be additive.
- For key analyses, unadjusted OR estimates will also be provided as sensitivity analyses.

The validity of the proportional odds assumption will be assessed by testing for heterogeneity in the log ORs (for the treatment effect) across the dichotomized cumulative ordered categories in the corresponding logistic regression model (partial proportional odds model, test for “unequal slopes”).

- The primary sensitivity analysis testing the proportional odds assumption will compare the unadjusted proportional odds model for the treatment comparison (null model) versus a partial proportional odds model that allows for “unequal slopes” across the dichotomized cumulative categories (i.e., when testing the proportional odds assumption for the treatment comparison with respect to the pulmonary outcome on Day 5, the model will allow for heterogeneous ORs across the Day 5 pulmonary categories) as well as across the stratification covariates (i.e., the baseline pulmonary+ categories and site pharmacy strata) (full partial proportional odds model).

**Continuous outcomes** will be compared between treatment groups using ANCOVA models for comparing means, if the ANCOVA model assumptions hold. If the distributions of the continuous outcomes are skewed, outcomes may be transformed, or compared between treatment groups using rank-based methods, such as the Wilcoxon test, or quantile (median) regression.

Comparisons between treatment groups for a continuous outcome will be adjusted for baseline values of the outcome, for the purpose of variance reduction, unless there are concerns over model stability with such an adjustment. For this purpose, the baseline value will be included as covariate in the model (e.g., ANCOVA, linear mixed models).

To estimate the treatment effect for longitudinally measured continuous outcomes, the outcome will usually be defined as “change from baseline” (difference at follow-up visit minus baseline value). The treatment effect through follow-up will then be estimated with 95% confidence intervals using generalized estimating equations (GEE) with an indicator for treatment group, or, in the case of Gaussian responses, the corresponding mixed effects models with random effects for participants. When there is more than one follow-up visit, “visit number” (day) may be included as categorical variable in the model, for variance reduction; alternatively, “time” may be included as continuous variable. Models will also be adjusted for the baseline values of the outcome variable.

**Time-to-event outcomes** will be summarized with Kaplan-Meier estimates for cumulative probabilities over time, and compared between treatment groups using log-rank tests or Cox proportional hazards models, or the corresponding competing risk analogues when death is a competing risk for the outcome. In particular, the primary endpoint of “time to sustained recovery” will be analyzed taking into account the competing risk of death. The following competing risk methods will be used:

- Aalen-Johannsen estimator for the cumulative incidence function (analogue to the Kaplan-Meier estimate)^3^
- Gray’s test with ρ=0 (analogue to the log-rank test)^4^
- Fine-Gray estimates and tests for the sub-distribution hazard ratio (analogue to the Cox proportional hazards model).^5,6^

The proportional hazards assumption will be tested by adding an interaction term for time by treatment group to the model. The cumulative proportions of participants who experienced the event will also be compared at given time points (specified in secondary objectives, e.g., at 28 days); in this case, the cumulative proportions will be estimated using Kaplan-Meier estimates or the competing risks analogue, and/or as proportion of participants who reached the time point (e.g., time since randomization ≥ 28 days).

The **administrative follow-up time** is defined as the minimum of (cut date minus randomization date) or the analysis time period. For example, the analysis time period for the primary endpoint of *sustained recovery* is 90 days, and the analysis time period for the important safety endpoint, the composite of *grade 3 and 4 events, SAEs, organ failure, serious infection or death*, is 5 days or 28 days. The **administrative censoring date** is the earlier of the cut-date of the dataset, or randomization date plus analysis time period.

**Comment:** The notion of “administrative censoring” is important in time-to-event analyses in the presence of competing risks. For example, the Fine-Gray method for estimating the sub-hazard ratio for sustained recovery can be approximated by using a Cox proportional hazards model where follow-up time for participants who died prior to achieving sustained recovery is not censored at death, but at the administrative censoring date.

**Censoring for time-to-event analyses**

For **interim** analyses, the type of censoring used will depend on the data collection schedule.

- If the reporting of the endpoint is data-driven (e.g., SAEs and deaths are reported as they occur), then follow-up is censored at the administrative censoring date, at the date of withdrawal, or loss to follow-up, whichever occurs earliest.
- If the date of the event is elicited retrospectively at fixed study visits spaced more than one week apart (e.g., “sustained recovery”), follow-up will be censored at the last day the endpoint status was ascertained.
- Sensitivity analyses will be provided for key analyses when the outcome status is uncertain.

For **final** analyses, follow-up will be censored on the last day the outcome status was ascertained.

**Adverse events** (AEs) will be classified by system organ class according to MedDRA®^[1]^ (currently version 23.1 [September 2020] is used; when new versions are implemented, items are recoded). AEs will be graded according to the *DAIDS Table for Grading the Severity of Adult and Pediatric Adverse Events*, *Corrected Version 2.1 (July 2017)* (also referred to as the *DAIDS AE Grading Table*).^7^ Cause of death will also be coded according to MedDRA®.

The number and percent of participants with grade 1-4 AEs will be summarized by day and grade, and by MedDRA® System Organ Class and grade. The percentage of participants with AEs will be compared between treatment groups according to grade cut-offs, e.g., “percent of participants with any AE”, “percent of participants with grade 2 or higher AEs”, etc., using CMH tests. The total number of events and median (IQR) of events per participant will also be summarized.

Additionally, the incidence of grade 3 and higher AEs will be summarized (number and percent of participants), and compared between treatment groups using time-to-event methods.

**Significance level, two-sided tests:** Unless noted otherwise, statistical tests and confidence intervals will be 2-sided, confidence intervals will have approximate 95% coverage probability, and test results with P-values ≤ 0.05 will be considered “significant”. Percentages will be reported to at least one decimal place. P-values will be given to 2 significant figures.

**Cut-date for interim reviews:** Analysis data sets will be frozen (locked) several days (or weeks) prior to the review date, to allow the unblinded statisticians time to prepare a consistent report. The cut-date may be earlier than the date of the data freeze, to allow for lag time in the reporting of events. Early in the trial, the cut date and freeze date will be very close to the review date, to ensure timely safety reviews.

### 4 Enrolment and Eligibility

For the open report, the following enrolment and eligibility summaries will be provided:

- Enrolment over calendar time: plot by day or week, cumulative and increments.
- Enrolment by site pharmacy and by country: number (%)
- Eligibility: number (%) and reasons for eligibility violations

These summaries will be provided overall, and by disease severity randomization stratum.

For the closed report, enrolment and eligibility violations will be summarized by treatment group.

### 5 Baseline Characteristics

Baseline characteristics will be based on information collected on baseline and screening forms. For the open report, baseline characteristics will be summarized pooled across the two treatment groups (investigational agent and the “pooled” control group as described in section 2 above).

For the closed report, baseline characteristics will be summarized by treatment group.

The following baseline characteristics will be reported; unless noted otherwise, categorical variables will be summarized with numbers (%) in each category, and continuous variables will be summarized with median (IQR); in the open report, in addition, the mean (SD) and range may be provided.

- Demographics
  - Age: distribution in categories 18-29, 30-39, 40-49, 50-59, 60-69, 70-79, ≥80 years; and summary as continuous variable
  - Sex at birth: number (%) male, female
  - Ethnic group: number (%) Asian, Black, Latino/Hispanic, White, other
  - Type of residence (“home”)
  - Country of enrolment
- COVID-19 related characteristics
  - Duration of symptoms prior to enrolment
  - Use of remdesivir prior to enrolment
  - Pulmonary and pulmonary+ ordinal outcomes, number (%) in each category
  - NEWS: summary as continuous variable
  - Respiratory function scale (modified Borg dyspnea scale; continuous outcome)
  - Disease severity randomization stratum (for investigational agents that enrol in both strata), number (%) in each category
  - Receipt of SARS-CoV-2 vaccination, and type of vaccine (and if received as part of a blinded clinical trial, in which case the vaccine may be active or control)
- Other clinical characteristics
  - Concomitant treatments
  - Corticosteroid use will be summarized overall, and separately by oxygen requirement at baseline (no supplemental oxygen, < 4 L/min, conventional supplemental oxygen ≥ 4 L/min, high-flow oxygen or mechanical ventilation/ECMO)
  - History of chronic conditions (cardiovascular disease, diabetes, asthma, chronic obstructive pulmonary disease, hypertension, chronic kidney disease, hepatic impairment, cancer, or immunosuppressive disorder [HIV, and other than HIV])
  - Prior cerebrovascular event
  - Prior myocardial infarction (MI)
  - Requirement of continuous chronic supplemental oxygen
  - BMI (<30, 30-39.9, 40+)
  - Pregnancy (not applicable to protocol versions 2 and 3)
- Laboratory values: as continuous outcomes, and number (%) of grade 3 or 4 abnormalities according to the *DAIDS AE Grading Table.*

Some biomarkers will be measured centrally from stored samples, for example, SARS-CoV-2 antibody levels and SARS-CoV-2 viral RNA. If these measures are available, they will be included in interim reports.

### 6 Administration of Study Treatment

These data are an important part of the safety review, with particular emphasis on infusion-related reactions and symptoms occurring during or within up to 2 hours after the infusion. These reactions and symptoms will be graded according to the DAIDS AE Grading Table.

The administration of study treatment is also an essential element of study conduct. Several summaries, pooled across treatment groups, will be included in the open report or provided to study leadership. Any summaries of adverse events or infusion-related reactions are restricted to the closed report.

**Each investigational agent is administered as a one-time infusion**. The following statistics will be used to summarize the infusion in each treatment group (active and control):

- Number and percentage of participants receiving complete infusion, partial infusion, infusion paused but resumed for complete infusion, or not infused (comparison by intention-to-treat).
- Number and percentage of participants with infusion-related reactions and symptoms (reported during the infusion or within 2 hours after the infusion), by grade. (Closed report only)
- Number and percentage of participants with an incident AE, SAE, UP or SUSAR on Day 0 during or after the infusion, overall and by oxygen requirement category at time of infusion (oxygen requirement at baseline, unless updated information is available; categories: no supplemental oxygen, < 4 L/min, conventional supplemental oxygen ≥ 4 L/min, high-flow oxygen or mechanical ventilation/ECMO). Types of AEs will be summarized by system organ class and by grade. (Closed report only)
- Number and percentage of participants who received:
  - Prior to infusion, medication to prevent infusion reactions, and type of medication
  - During or within 2 hours after infusion, medication to treat infusion reactions, and type of medication (Closed report only)
- Among participants infused, the day of infusion (same day as randomization, next day, > 1 day after randomization), and time between randomization and beginning of infusion (median hours, IQR).
- Among participants receiving full infusion, duration of infusion (median minutes, IQR).
- Time from vial puncture (beginning of preparation of the study agent by the pharmacist) to the end of the infusion, and number and percent of participants for whom the agent-specific time window was exceeded.
- Remdesivir:
  - Number and percent of participants who received (any) remdesivir, and number of days remdesivir was administered: median, IQR, distribution. (Closed report only)
  - Number and percent of participants who received remdesivir prior to the day of randomization, overall and by number of days prior
  - On the day of randomization: Number and percent of participants who received remdesivir prior to the investigational agent; after the investigational agent; no remdesivir.

Treatment groups will be compared by mITT (excluding participants who did not receive any investigational agent/placebo), unless specified otherwise. The treatment comparisons will be performed using the methods described in section 3 for binary and continuous outcomes (stratified CMH test for comparing percentages, and Wilcoxon rank-sum test [or quantile regression for comparing medians], respectively).

Selected summaries will also be provided separately for the two disease severity strata.

**Analyses specific to BRII-196 and BRII-198**

The investigational agent by Brii Bio consists of two nMAbs (BRII-196 and BRII-198) that are administered as consecutive one-time IV infusions.

- The infusion status,
  - complete infusion, partial infusion, or not infused
  - for those where infusion was paused for adverse reaction, whether the infusion was later completed,

will be summarized overall, and separately for BRII-196 and BRII-198. The order of infusion for BRII-196 and BRII-198 will be described.

### 7 Safety Analyses

The planned timing of safety reviews is described in section 2. An overview of the safety data collection is provided in Appendix C.

**Analysis cohort:** Safety analyses will be carried out on participants who received a complete or partial infusion of the investigational agent (modified intention-to-treat [mITT]), unless otherwise stated.

A comprehensive safety review includes:

- Comparison of the treatment groups for the primary safety endpoint, its components, and analyses of secondary safety outcomes (described in this section)
- Analyses of infusion-related reactions and symptoms, described in section 6
- Evaluation of the “efficacy outcomes” (the pulmonary and pulmonary+ ordinal outcomes early in follow-up, and time to sustained recovery), which contain important safety information.

In addition to the full DSMB reviews, more frequent, shorter safety reports may be provided to the DSMB, for example, weekly safety reports early in the trial.

This section describes the primary safety outcome, and the analyses of AEs, SAEs, UPs, SUSARs, and deaths. Comparisons between treatment groups will be stratified by study pharmacy and by disease severity at study entry (as described in section 3 under “stratification”).

In order to streamline the reporting of events, it was decided that certain protocol-specified exempt events (PSEE) are *not reported as SAEs*, unless they are considered related to the study treatment by the investigator. While the PSEEs in this protocol are similar in severity to SAEs, PSEEs are reported not on the SAE eCRF, but are reported as study endpoints on various other eCRFs. The *clinical organ failure or serious infections* composite outcome, described in Appendix B, is the composite of all PSEEs.

The following safety and tolerability outcomes will be analyzed; models will be stratified by disease severity and study site pharmacy, as described in section 3 under “stratification”, unless noted otherwise:

- The **primary safety endpoint** is a composite of incident grade 3 or 4 clinical adverse events, SAEs, clinical organ failure or serious infections (PSEE), or death through Day 5. The number and proportion of participants experiencing one of these events up through Day 5 will be tabulated, and treatment groups will be compared using a CMH test stratified by study site pharmacy and by disease severity at study entry.
  - Mortality will be analyzed as a key secondary outcome, see below.
  - The individual components of the composite outcome will be summarized.
  - Sensitivity analyses for the primary safety outcome: For interim analyses, while the trial is still enrolling, treatment groups will also be compared for time to event through Day 5 using a log-rank test, stratified by site pharmacy and disease severity at study entry; the HR will be estimated with a 95% CI using a Cox proportional hazards model, and the cumulative proportion of participants with events over the first 5 days in each treatment group will be estimated using Kaplan-Meier curves.
- All-cause mortality through follow-up will be analyzed using time-to-event methods. Cumulative proportions of participants who died in each treatment group will be estimated using Kaplan-Meier estimates, and summarized in tables (proportion of participants who died by Days 5, 7, 14, 28, 60, 90, month 6, 12, and 18) and figures (Kaplan-Meier curves with pointwise 95% CIs). Treatment groups will be compared for time to death using log-rank tests, stratified by study site pharmacy and disease severity, and an overall HR will be estimated with 95% CIs using stratified Cox proportional hazards models.
- Cause of death will be MedDRA® coded and summarized by treatment group.
- The following composite endpoints will be analyzed using time-to-event methods (cumulative proportions of participants with events will be estimated using Kaplan-Meier curves with pointwise 95% CIs; treatment groups will be compared using log-rank tests; numbers and percent of participants with events will be summarized by treatment group, and overall HRs with 95% CI will be estimated using Cox proportional hazards models):
  - Composite of incident grade 3 or 4 clinical adverse events, SAEs, clinical organ failure or serious infection, or death through Day 28
    - Components of the composite endpoint will be also be summarized, overall and by system organ class. Proportions of participants who experienced any of these events by Day 28 will be compared using stratified CMH tests and logistic regression.
  - Composite of SAEs, clinical organ failure, serious infection, or death through Day 28 and Day 90
  - Composite of hospital re-admission or death through 18 months
- Treatment groups will be compared for the incidence of non-pulmonary events in the pulmonary+ ordinal outcome that are not part of the pulmonary outcome, through Day 5 and Day 7 (using time-to-event methods, with death as competing risk). These events are shown in red in Appendix A.
- AEs, SAEs, and UPs will be classified by MedDRA® system organ class. AEs will be graded for severity according to the *DAIDS AE Grading Table*. Grade 1-4 clinical AEs will be reported at baseline (Day 0 prior to infusion of the investigational agent), Day 0 after the infusion, Days 1-7, and on Days 14 and 28.

The number and percent of participants with AEs will be summarized by day (Day 0 separately prior and after the infusion) and grade, and by system organ class and grade. Comparisons between treatment groups will be for the proportion of participants with AEs of a given grade or higher (i.e., any grade, grade 2+, grade 3+, grade 4). The treatment comparisons will be performed using stratified CMH tests or logistic regression.

- For comparisons by day, the proportion of participants with any grade AEs will be compared for Days 0 (after infusion) through 7, and on Days 14 and 28.
- For comparisons by grade, the proportion of participants who experienced any grade AEs (grade 2+ AEs, etc.) between Day 0 (after the infusion) through Day 7 will be compared.
- For the comparison by system organ class, CMH tests will be performed if the number of participants with AEs is sufficiently large. System organ classes may be split up into MedDRA® preferred terms (PT) for system organ classes where the treatment difference is significant.

Other clinically meaningful AE groupings (beyond system organ class) may be developed by the study team, who are blinded to the treatment effect.

- In addition to any grade AEs through Day 7, grade 3 and 4 clinical AEs are being reported through Day 28. The number and percent of participants with incident grade 3 or 4 AEs through Day 28 will be summarized, overall and by system organ class, and compared between treatment groups using stratified CMH tests. (A grade 3 or 4 AE is considered “incident” if the event was not present at baseline or increased to grade 3 or 4 from grades 1 or 2.)

To illustrate the time course, the incidence of *grade 3 or 4 AEs or death* through Day 28 will be summarized by treatment group using Kaplan-Meier estimates of the cumulative incidence functions (CIF).

- Infusion-related reactions and symptoms during infusion or within 2 hours after infusion of the investigational agent or placebo, and infusion cessation prior to completion will be tabulated and compared between treatment groups; analyses are described in section 6.
- Treatment groups will be compared for the proportion of participants who developed *organ failure or serious infections* through Day 28 and through Day 90, overall and by individual components, using stratified CMH tests. Individual components of this composite outcome will be tabulated.
- Subgroup analyses: The impact of study arm on the primary safety outcome (composite of grade 3 or 4 events, SAEs, clinical organ failure, serious infections, or death through Day 5) and other important safety outcomes will be assessed for subgroups defined by baseline characteristics, including demographics, duration of symptoms at enrollment, baseline classification of “home”, clinical history and presentation (including disease severity stratum and pulmonary+ ordinal outcome at baseline), and tests for homogeneity of the treatment effect across subgroups will be carried out. Outcomes and methods for subgroup analyses are described in detail in section 8.4.
- Treatment groups will be compared for incidence of a composite of cardiovascular and thromboembolic events, a subset of the organ failure outcome (items 6b1, 6e2, 6e3, and 6f2 in Appendix B). Time-to-event methods will be used that take into account the competing risk of death (as described in section 3, using Aalen-Johansen estimates for the cumulative incidence functions, and Gray’s and Fine-Gray’s methods to compare treatment groups and estimate the sub-hazard ratio).
- Treatment groups will be compared for mean changes in laboratory test values from baseline to Day 5, and for incidence of grade 3 and 4 laboratory abnormalities at Day 5 (new abnormality or increase in grade). Laboratory tests are conducted locally, and include serum creatinine, AST/SGOT or ALT/SGPT, WBC, hemoglobin, platelet counts, lymphocyte counts, and C-reactive protein. Statistical methods are described in section 3.
- Participants who are pregnant are not eligible for enrolment. For participants who become pregnant, pregnancy outcomes will be summarized.
- In addition to the safety outcomes specified in the platform protocol, other targeted safety outcomes for specific investigational agents may be specified in appendices to the protocol. Analyses will be specified in the corresponding agent-specific appendix to this SAP.

Listings of SAEs, clinical organ failure and serious infections (PSEE), incident grade 3 and 4 AEs, UPs, SUSARs, and deaths (with cause of death) by treatment group will be provided at each DSMB meeting, with new events highlighted. The listings will include important baseline characteristics, such as age, sex, and disease severity (pulmonary outcome category) at study entry.

Further safety assessments may be considered.

Corticosteroid use will be monitored; concomitant medication use is collected at baseline and at Day 5.

- Corticosteroid use (any use at baseline or Day 5) will be summarized by treatment group and by oxygen requirement (worst category through Day 5: no supplemental oxygen,
  < 4 L/min, conventional supplemental oxygen ≥ 4 L/min, high-flow oxygen or mechanical ventilation/ECMO).
- Corticosteroid use on Day 5 will be summarized by treatment group and by oxygen requirement (worst category on Day 5, as above).

### 8 Efficacy Analyses

**Analysis cohort:** Comparisons between each investigational agent and its concurrently randomized (pooled) controls will be by intention-to-treat (ITT) unless otherwise stated.

#### 8.1 Primary Efficacy Endpoint and Primary Analysis

The **primary efficacy outcome** of the trial is “time from randomization to *sustained recovery* through Day 90”*. Sustained recovery* is defined as being discharged from the index hospitalization, followed by being alive and *home* for 14 consecutive days.

**Comment:** The shortest possible time to sustained recovery is 14 days (this would require the patient to be discharged from the hospital on the day of randomization), and a patient would have to be discharged from the index hospitalization no later than Day 76 to achieve *sustained recovery* by Day 90.

**Definition of *Home* for the primary endpoint:**

According to the protocol, section 4.2, *Home* is defined as the level of residence or facility where the participant was residing prior to hospital admission leading to enrollment in this protocol.

Residence or facility groupings to define home are:

1) **Independent/community dwelling** with or without help, including house, apartment, undomiciled/homeless, shelter, or hotel

2) **Residential care facility** (e.g., assisted living facility, group home, other non-medical institutional setting)

3) **Other healthcare facility** (e.g., skilled nursing facility, acute rehab facility)

4) **Long-term acute care hospital** (hospital aimed at providing intensive, longer term acute care services, often for more than 28 days).

Lower (less intensive) level of residence or facility will also be considered as home. By definition, “home” cannot be a “short-term acute care” facility. Participants previously affiliated with a “long-term acute care” hospital recover when they return to the same or lower level of care.

Readmission from “home” (to a higher level of care) may occur and if this occurs within 14 days of the first discharge to “home”, then the primary endpoint will not be reached until such time as the participant has been at home for 14 consecutive days.

Participants residing in a facility solely for public health or quarantine purposes will be considered as residing in the lowest level of required residence had these public health measures not been instated.

**Primary analysis**

The investigational agent will be compared to the (pooled) control group for *time to sustained recovery through Day 90* by intention-to-treat, using Gray’s test with ρ=0.^4^ The test will be stratified by disease severity at entry and by site pharmacy (as described in section 3 under “stratification”). Gray’s test compares the cumulative incidence functions for *sustained recovery* between the treatment groups, taking into account the “competing risk” of death in analyzing *sustained recovery*. Gray’s test with ρ=0 is the competing-risks analogue of the log-rank test.

**Comment:** Comparisons will be presented such that recovery rate ratios (RRR) >1 denote superiority of the investigational agent.

Analyses for the *sustained recovery* endpoint require methods that take into account the competing risk of death, as participants may die before ever achieving *sustained recovery*. The *sustained recovery* outcome requires knowledge of a participant’s residence status for at least 14 days after arriving “home” (as defined above).

- The cumulative incidence functions for sustained recovery will be estimated by treatment group, using Aalen-Johansen estimators.^3^ The estimates will be plotted over time, and tabulated at selected time points (days 15, 21, 28, 42, 60, 75, 90). The Aalen-Johansen estimator for a cumulative incidence function is the analogue of the Kaplan-Meier estimator in the presence of competing risks.
- The recovery rate ratio (RRR) for time to sustained recovery of the investigational agent versus control will be estimated, as a point estimate with a 95% CI, using the Fine-Gray model, stratified by disease severity at study entry and study site pharmacy.^5,6^ The corresponding p-value for RRR=1 versus the two-sided alternative will be calculated. The Fine-Gray method is the competing risks equivalent of Cox proportional hazards models; the RRR compares the cumulative incidence rates of *sustained recovery* between the study arms and is a sub-distribution hazards ratio.
- To aid in the interpretation of the estimated treatment difference, the median days to sustained recovery (through Day 90) will be estimated for the investigational agent and the control group. Medians will be compared using the Wilcoxon rank sum test or quantile (median) regression. Participants who die at any time up to Day 90 will be assigned 91 days.

**Censoring:**

- Participants who are alive but have not experienced sustained recovery will be censored at the last date the endpoint status was ascertained (for interim analyses as well as the final analysis).
- For interim monitoring, two sensitivity analyses will be performed:
  1. Follow-up for time to sustained recovery will be censored *administratively* at the cut-date for the current report or Day 90, whichever comes first, with last known endpoint status carried forward. For participants who died, this type of censoring is integrated into Gray’s test; using a log-rank test would require carrying forward the “not recovered” status for participants who died, up to the administrative censoring date.
  2. Administrative censoring as described above will be applied, with the modification that participants who have been discharged from the hospital, were “home” at the latest date when residence was ascertained (but for < 14 days), and, if they have remained there, would have been at home for 14+ days by the cut-date, will be imputed as having experienced *sustained recovery* (achieved on day 14 at home).

Participants who withdrew consent or were lost to follow-up will be censored at the date of withdrawal or the last date the endpoint status was known, respectively.

In the first sensitivity analysis, the “not recovered” status is carried forward to the administrative censoring date; in the second analysis, “sustained recovery” is assumed at the earliest possible date. The first analysis potentially underestimates the rate of recovery, whereas the second analysis overestimates the recovery rate. In all analyses for time to sustained recovery, death is treated as competing risk.

**Ascertainment of sustained recovery**

The date of discharge from the index hospital will be recorded. Irrespective of the timing of the hospital discharge, there will be patient contact approximately every two weeks, on Days 14, 28, 42, 60, 75, and 90, either at a scheduled clinic visit or through phone contact. At these time points, a) vital status, and b) the location of the participant over time will be recorded, to assess whether the participant had been “at home” for 14 days. Therefore, the outcome status of sustained recovery will usually be ascertained within 3 weeks or less of the date the outcome was achieved.

- To illustrate the status of the primary endpoint, the recovery status of participants will be described over time with the following categories (at interim reviews):
  1. At home for 14+ days (reached the primary endpoint of sustained recovery)

*Did not reach sustained recovery, and:*

2. At home, < 14 days

3. Discharged from the hospital, but not at home

4. Hospitalized

5. Dead

6. Primary endpoint status unknown.

The proportions of participants in each of the 6 categories will be summarized over time, by treatment group (stacked bar graphs and tables). In this analysis, both “sustained recovery” and “death” are absorbing states.

**Assessment of model assumptions**

- The trial was powered to detect an RRR of 1.25 with 90% power; this requires 843 sustained recoveries among the 1000 participants by Day 90. The rate of recoveries will be monitored, overall and within the two disease severity strata. Deviations of the observed distribution from the hypothesized distribution in the control arm will be monitored, and the impact on the power of the trial will be assessed. Prior to the completion of the trial for the investigational agent, sample size will be re-estimated by the blinded statisticians on the study team, based on the pooled rate of *sustained recovery*.
- The Fine-Gray model assumes that the sub-distribution rate ratio for *sustained recovery* is constant over time, similar to Cox proportional hazards models. The assumption of constant RRR will be tested by including an interaction effect between time and treatment indicator.

**Sensitivity Analyses**

- As sensitivity analyses, the primary comparison will be repeated after excluding participants who did not receive any of the investigational agent/placebo (modified intention-to-treat).
- Sensitivity analyses for the primary endpoint comparisons will include consideration of home oxygen above pre-morbid oxygen use (described in section 4.2.2 item 12 of the protocol). For these sensitivity analyses, *sustained recovery* will be re-defined as:
  a. “Discharged to home, alive at home without use of continuous supplemental oxygen for an uninterrupted 14 day period”
  b. “Discharged to home, alive at home for an uninterrupted 14 day period, and no supplemental oxygen use a the end of the 14 day period”
- If the RRR is not constant (test described under “assessment of model assumptions” above), as a sensitivity analysis, the RRR will be estimated within time periods, for example, Day 14-28, Day 29-60, Day 61-90.
- Additional sensitivity analyses are described under “censoring” above.

#### 8.2 Key Secondary Outcomes

- Mortality is a key secondary outcome; analyses are described in section 7.
- To supplement the separate analyses of time to sustained recovery and time to death, the two endpoints will be analyzed jointly using the “win ratio” method for the composite outcome of time to recovery or death.^8^ At a given time point (Day 90), the win ratio statistic ranks participants’ outcomes into three ordered categories, death, alive but not achieved sustained recovery, alive and achieved sustained recovery; ties are broken by time since randomization. So, time to death is first used to determine the winning group (i.e., longer time to death), then time to sustained recovery is used to determine the winning group (i.e., shorter time to recovery): in this manner these conflicting outcomes can be combined into a composite while recognizing the importance of mortality. Matching on baseline disease severity will be used to estimate the win ratio statistic. This combination of time to sustained recovery and time to death is also a key secondary analysis.

#### 8.3 Other Secondary Outcomes

The protocol defines a number of secondary endpoints in addition to the two key endpoints described in section 8.2 above. These analyses will be carried out for the final report. Selected secondary endpoints may also be analyzed for interim monitoring reports, to help evaluate the safety and efficacy of the investigational agent.

Below, the secondary outcomes from section 4.2.2 of the protocol are cited, with a short description of the analysis methods. For each outcome, the treatment groups will be compared by intention-to-treat, stratified by disease severity at study entry and by site pharmacy, as described in section 3 under “stratification”.

- Time to discharge for the initial hospitalization. Treatment groups will be compared using time-to-event methods that take into account the competing risk of death, similar to the analyses for time to sustained recovery described in section 8.1.
  - Hospital readmissions will be summarized using methods for recurrent events (i.e. those who are readmitted will re-enter the risk set).^9^
- Days alive outside of a short-term acute care hospital up to day 90. For this analysis, the “last-off” method will be used, i.e., days from the latest hospital discharge to day 90 will be counted. A person who dies within 90 days will be assigned a value of 0, consistent with the approach taken in trials of intensive care-based interventions. We will present the median days by group and test the hypothesis of no difference between arms with a Wilcoxon rank sum test.
  - For interim analyses, only participants who have reached Day 90 (administrative follow-up for those who died) will be included, to avoid bias. Alternatively, a shorter time period may be used.
- Pulmonary+ and pulmonary ordinal outcomes on Days 1-7, and the pulmonary ordinal outcome on Days 14 and 28. The proportion of participants in each category of the pulmonary and pulmonary+ outcomes will be summarized over time (both outcomes at days 1-7, the pulmonary outcome also at Days 14 and 28); at each of those days, treatment groups will be compared using proportional odds models as described in section 9.1; the proportional odds models will be adjusted for the categories of the pulmonary+ outcome at baseline and for study site pharmacy. If participants in both disease severity strata are enrolled, the models will also be adjusted for the interaction between disease severity stratum and pharmacy.

Additionally, the ordinal outcomes will be dichotomized (“category 1”, “best 2 categories” through “best 5 categories”), and proportions will be compared between treatment groups at selected time points using logistic regression. For these analyses, the key dichotomized outcome considers the “best 2 categories”, which is similar to the “recovery” outcome in the ACTT-1 trial.

- *Clinical organ failure or serious infections*, defined by development of any one or more of the clinical events listed in Appendix B, through Days 28 and 90. The development of *organ failure or serious infections* will be analyzed as a binary outcome, and the proportions of participants who developed *organ failure, serious infections or death* will be compared across arms using stratified CMH tests, overall and for individual components.
- A composite of death, clinical organ failure, or serious infection through Days 28 and 90 (see Appendix B). Treatment groups will be compared using standard time-to-event methods, since death is part of the outcome and not a competing risk. (Also described in section 7 as safety analysis).
- Outcomes assessed in other treatment trials of COVID-19 for hospitalized participants in order to facilitate cross-trial comparisons and overviews (e.g. 6- , 7-, and 8-category ordinal scales assessed at Days 1-7, 14 and 28; time to improvement in 1 or 2 categories of ordinal scale; time to best 3 categories of ordinal scale, and binary outcomes defined by improvement or worsening based on other ordinal outcomes). We will try to match the analyses in the other trials, to get results that can be compared. These analyses will not be performed for interim reports to the DSMB, unless requested.
- A composite of cardiovascular events (outcomes listed in items b1, e2 and e3 in Appendix B) and thromboembolic events (item f2). Time to event methods will be used that take into account the competing risk of death, e.g., Gray’s test to compare treatment groups.

#### 8.4 Subgroup Analyses

As stated in the protocol, subgroup analyses for the primary efficacy outcome (time to sustained recovery), the primary safety outcomes (composite of grade 3 and 4 events, SAEs, clinical organ failure, serious infections, or death through Day 5 and Day 28, composite of SAEs, clinical organ failure, serious infections, or death through Day 90), and for time to death will be performed to determine whether and how the treatment effect (active versus control) differs qualitatively across various subgroups defined at baseline, and whether there are safety concerns in specific subgroups.

Subgroup analyses will also be carried out for the Pulmonary and Pulmonary+ ordinal outcomes on Day 5.

Key subgroup analysis are by disease severity and by the categories of the pulmonary+ outcome at baseline; other important subgroups include subgroups by duration of symptoms prior to enrollment, by age and by pre-existing conditions.

Subgroup analyses will be performed by the following baseline factors:

- Disease severity (categories of the pulmonary+ outcome at study entry, randomization stratum). This subgroup analysis will be used at interim analyses after expansion of enrollment to assess if the treatment effect varies across the severity strata.
- Duration of symptoms prior to enrollment
- Age (18-49, 50-59, 60-69, 70-79, 80+)
- Biological sex
- Race/ethnicity
- Geographic location
- Residence (home) at the time COVID-19 symptoms developed
- Body mass index (BMI)
- History of chronic conditions (cardiovascular disease, diabetes, asthma, chronic obstructive pulmonary disease, hypertension, chronic kidney disease, hepatic impairment, or cancer)
- SARS-CoV-2 vaccination status at baseline.

When SARS-CoV-2 viral load, antibody, and antigen levels are available, subgroups will also be considered by upper respiratory SARS-CoV-2 viral load, by antibody level, by neutralizing antibody level, and by antigen level at baseline. The lab tests to be conducted and the analysis plan for these endpoints are currently under discussion.

Subgroup analyses for the primary endpoint of time to sustained recovery will use the Fine-Gray model, stratified by disease severity at study entry, and by site pharmacy if the sample size permits. RRRs with 95% CIs comparing the investigational agent versus control will be estimated for each subgroup. Global tests for heterogeneity of the treatment effect across subgroups will be carried out, by adding the interaction between the subgroup indicator and the treatment group indicator to the model. In case the subgroup was formed by categorizing a continuous variable, the interaction term will be formed between the subgroup indicator and the continuous variable.

Subgroup analyses for the primary safety endpoint at Day 5 will use logistic regression, stratified by disease severity at study entry, and by site pharmacy (if the sample size permits). Subgroup analyses for safety endpoints that are analyzed using time-to-event methods (those analyzed through Day 28 or longer) will use stratified Cox proportional hazards models, since death is part of the composite endpoints and not a competing risk. HRs will be estimated for each subgroup, and global tests of heterogeneity of the treatment effect will be carried out, as described above.

Subgroup analyses for the pulmonary and pulmonary+ ordinal outcomes on Day 5 will use proportional odds models, adjusted for pulmonary+ category at study entry and for site pharmacy.

Additionally, subgroup analyses will be conducted for subgroups formed by a disease progression risk score at baseline. The construction of this risk score will be revisited as data on the sustained recovery endpoint accumulate for new investigational agents.

Subgroup analyses will not be adjusted for multiple comparisons. Subgroup analyses will be interpreted with caution due to limited power and uncontrolled type I error.

### 9 Interim Monitoring Guidelines for the DSMB

Each investigational agent versus placebo comparison will be treated as a separate clinical trial; stopping boundaries will be derived to allow for multiple interim looks, but will not be additionally inflated to adjust for simultaneous analysis of multiple investigational agents, except when explicitly stated in the agent-specific protocol appendix and statistical analysis plan.

The DSMB will be asked to recommend early termination or modification only when there is clear and substantial evidence of a treatment difference (unless a trial is stopped for futility).

#### 9.1 Early Assessment of Safety

For investigational agents with minimal pre-existing data, the pace of enrollment will be initially restricted and the DSMB will be asked to review safety data for the first 20 to 30 participants before increasing the pace of enrollment.

Subsequently, the DSMB will carry out regular reviews of safety data reports. These reports will include summaries of infusion–related events, grade 1-4 AEs, SAEs, organ failure, serious infections, and deaths, including the primary safety outcome at Day 5, and Day 28. Event listings for incident grade 3 and 4 AEs, SAEs, organ failure and serious infections (PSEE), SUSARs, UPs and deaths will be provided (events that were reported since the previous review will be marked). Narratives will be provided for selected SAEs, SUSARs or UPs, particularly those judged related to study treatment. Analyses are described in section 7.

**Monitoring boundary for harm:**

- Until the first 300 participants are enrolled (150 per study arm) and the early futility analysis is conducted, the treatment groups will also be compared for the “pulmonary” and “pulmonary+” ordinal outcomes at Day 5, using a proportional odds model stratified by study pharmacy and pulmonary+ outcome category at baseline. A Haybittle-Peto boundary with 2.5 standard deviation (SD) for the first 50 participants enrolled and 2.0 SD afterwards will be used as a guideline for harm.
- After the study population is expanded to include disease severity stratum 2, these analyses are performed by disease severity stratum.

At the discretion of the DSMB, these safety reports will be prepared at a frequency they specify, for example, weekly. The DSMB may also request additional data summaries.

#### 9.2 Early Assessment of Futility

##### 9.2.1 Interim Monitoring Guidelines for Early Assessment of Futility

Early in the trial, enrolment is restricted to participants in disease severity stratum 1. When Day 5 data for the first (approximately) 300 participants (150 per study arm) are available, the DSMB will review interim data and use pre-specified guidelines for early evidence of sufficient activity of the investigational agent that justifies continuing enrolment for the agent and expanding eligibility criteria to include participants in disease severity stratum 2 as well as stratum 1.

The early futility monitoring uses two co-primary outcomes, denoted by “pulmonary” and “pulmonary+”, assessed on Day 5. Both are ordered categorical outcomes with 7 categories, described in section 4.1 of the protocol and in Appendix A to this SAP. The pulmonary outcome considers largely respiratory-related disease, similar to the ordinal outcome in the ACTT-1 trial.^10^ The pulmonary+ outcome has the same categories for pulmonary complications (e.g., requirements for oxygen), and additionally includes extra-pulmonary outcomes such as thrombotic, myocardial, and cerebral complications of COVID-19.

Guidelines for the early futility assessment are as follows:

a. If the investigational agent is superior to placebo (i.e., p ≤ 0.3 for a one-sided test) in both the pulmonary+ and pulmonary intermediate ordinal outcomes, then enrolment for the agent will expand to complete the trial.
b. If there is insufficient evidence for superiority versus control (i.e., one-sided p>0.3) in each of the two outcomes, then stop randomization.
c. If there is evidence (1-sided p < 0.3) for an association for one endpoint and not the other, then the agent may or may not advance depending on the risk/benefit profile emerging from the data at this early stage. If the effect estimate for both outcomes is on the side of benefit, the preference would be towards advancing the agent and expanding enrollment to include disease severity stratum 2.

The DSMB will be asked to review whether the discordance is attributable to a positive or negative effect on extra-pulmonary organ dysfunction (the difference in the two ordinal scale categories, the conditions included in pulmonary+ but not in the pulmonary endpoint), and whether the same ordinal outcomes assessed on other days yield similar results, and weigh the risk/benefit profile. For example, if there is a significant positive effect on the pulmonary score and the lack of significant effect on the pulmonary+ score is driven by a lack of difference in the milder thrombotic symptoms in category 4 of the pulmonary+ scale (e.g. deep venous thrombosis) and there is no evidence of any raised risk of thrombosis overall, the agent will advance. Conversely, if the agent is superior to the control group with respect to the pulmonary outcome, but clearly inferior to the control group with respect to the pulmonary+ outcome or has a concerning safety profile, it will not advance.

Analyses of the primary efficacy endpoint, time to sustained recovery, will also be provided to the DSMB, as supporting information. These analyses are described in section 8.1.

After considering the aforementioned guidelines, the DSMB will be asked to consult with the Food Drug Administration before making their recommendation in order to consider any relevant external information.

##### 9.2.2 Analyses of the Pulmonary and Pulmonary+ outcomes on Day 5

- Treatment groups will be compared by intention to treat.
- For each of the two ordinal outcomes, the number and percentage of participants in each of the categories on Day 5 will be tabulated, and the OR of the active versus control group will be estimated using a proportional odds model with indicators for the investigational agent group (active versus control) and for the categories of the ordinal pulmonary+ outcome at baseline (to adjust for baseline severity of illness).^2^ The model will be stratified by site pharmacy.

The summary tables will show the adjusted summary OR with 95% CI, estimated as described above, as primary analysis. In addition, the unadjusted summary OR with 95% CI will be shown as sensitivity analysis (estimated using a proportional odds model without adjustment for the pulmonary+ baseline category or site pharmacy). In the case that the adjusted OR differs substantially from the unadjusted OR, the reason for the deviation will be explored.

**Comments:**

- Results will be presented such that OR>1 favors the investigational agent, denoting higher odds of more favorable disease categories in the group randomized to investigational agent compared with control.
- In order to avoid overestimating the proportion of participants who died, participants who died prior to Day 5 will only be included in the Day 5 summaries of the pulmonary and pulmonary+ outcomes if their time from randomization to cut-date is at least 5 days, and similarly for analyses on other days. Mortality is a key secondary endpoint and will be summarized cumulatively as an additional analysis.
- **For the initial futility analysis**, the tests comparing the investigational agent versus placebo will be performed using a (1-sided) type 1 error rate of 30%. This means, the investigational agent will be considered “superior” to the control with respect to the pulmonary (or pulmonary+) outcome, if the estimated summary OR is greater than 1, and the p-value ≤ 0.30.

The summary reports will show the estimated summary OR with 95% CI, the signed Z-value for the test statistic comparing the treatment groups, and the one-sided p-value for superiority, calculated in the primary analysis (i.e., using the proportional odds model that is adjusted for the pulmonary+ category at baseline and stratified by site pharmacy, as described above). As sensitivity analysis, these values will also be calculated using the unadjusted proportional odds model and included in the summary report.

**Comment:** At the recommendation of the FDA, patients requiring high-flow oxygen or mechanical ventilation (invasive or non-invasive) are currently not eligible for enrolment. Also, prior to the initial futility assessment, eligibility is restricted to disease severity stratum 1. Therefore, adjusting the treatment comparison for the pulmonary+ outcome at baseline is identical to adjusting for the following categories defined by oxygen requirement:

- No supplemental oxygen (pulmonary+ category 2)
- Supplemental oxygen < 4 L/min (or < 4 L/min above premorbid requirements) (pulmonary+ category 3)
- Supplemental oxygen ≥ 4 L/min (or ≥ 4 L/min above premorbid requirements, but not high-flow oxygen) (pulmonary+ category 4)
- To supplement the overall summary odds ratios for the 7-category outcomes, each dichotomized definition of improvement that can be formulated from the components of the ordinal outcomes will be considered separately; for example, treatment groups will be compared for the proportions of participants in category 1 on Day 5, proportions in categories 1 or 2 (“best two categories”), in categories 1-3, etc. Proportions will be tabulated, and odds ratios for active versus control groups will be estimated with 2-sided 95% CIs using logistic regression models. These analyses need to be interpreted with caution, because they are not adjusted for inflation of type I error due to multiple comparisons.
- Subgroup analyses will be carried out for the Pulmonary and Pulmonary+ outcomes on Day 5, to supplement the early futility analyses. The goal is to determine whether the treatment effect differs across subgroups, and to aid the DSMB in considerations on whether there are safety concerns in specific subgroups. Principles for subgroup analyses are described in section 8.4; here, subgroup analyses are based on the proportional odds models. In particular, heterogeneity of the treatment effect across the baseline pulmonary+ categories will be assessed.
- After an investigational agent has passed initial futility assessment and enrollment has been expanded to include participants in disease severity stratum 2, treatment comparisons for the pulmonary and pulmonary+ outcomes on Day 5 continue, and will be performed separately for each of the two disease severity strata, to assess safety for the more severely ill participants in stratum 2.

**Missing data: Unknown outcome status for the pulmonary or pulmonary+ outcomes on Day 5:**

The following items describe how missing data will be treated for the primary analyses of the pulmonary or pulmonary+ outcomes on Day 5. As needed, these methods may be also applied to analyses at other time points (e.g., Day 7).

- **Interim analyses**:
  - Only participants with Day 5 data for the pulmonary outcome will be included for the Day 5 comparisons. The number and proportion of participants with unknown outcome status will be summarized.

**Comment:** If the cut date is less than 10 days before the data freeze date, Day 5 data for the ordinal outcomes are considered “missing” only for participants with at least 10 days of administrative follow-up.

- **Final analyses** after completion of the trial:
  - If Day 5 data are missing for a substantial proportion of participants (e.g., more than 5%), multiple imputation will be used to impute missing Day 5 data for the pulmonary and pulmonary+ outcomes. For the imputation, the following baseline covariates will be considered in addition to the indicator for treatment group: age, sex, country, duration of symptoms prior to enrollment, status of the ordinal pulmonary (or pulmonary+) outcome, and presence of comorbidities. Ten rounds of imputation will be used to estimate the summary odds ratio.
  - The number and proportion of participants with missing data will be reported.

**Sensitivity analyses**

- As sensitivity analyses, the treatment groups will be compared by **modified intention-to-treat (mITT)** after excluding participants who did not receive any of the assigned investigational agent (active or control). This mITT analysis will be provided at important decision points, e.g., when the test statistic approaches the monitoring boundary, and for the final analyses after completion of the trial.
- Treatment groups will be compared for the pulmonary and pulmonary+ outcomes on Days 1-7, to monitor the consistency of the treatment effect over time.

**Assessment of model assumptions**

- For the pulmonary and pulmonary+ outcomes at Day 5, the proportionality assumption of the odds ratio will be assessed (by including the interaction between the treatment group indicator and indicators for the Day 5 cumulative ordinal categories in the model, as well as the interactions between the treatment group indicator and the indicators for the strata by baseline pulmonary+ categories and site pharmacy; this corresponds to testing for separate slopes using a partial proportional odds model, see section 3 under “ordered categories”). If there is evidence for non-proportionality, the summary odds ratio in the proportional odds model will still be used to quantify the treatment effect, and the analyses of the dichotomized ordinal outcome categories will be used to help interpret the treatment effect.
- The sample size of 300 (150 per study arm) is sufficient to detect a summary OR of 1.60 for the comparison of the investigational agent versus control for each of the two ordinal outcomes with 95% power. The power of the tests depends on the hypothesized OR and the hypothesized distribution in the control group used for the sample size calculations. At the time of the early futility review, the deviation of the observed distribution from the hypothesized distribution in the control arm will be assessed, and the impact on the power of the trial will be estimated.

#### 9.3 Interim Monitoring Guidelines for the Primary Endpoint

This section describes the interim monitoring guidelines that apply after the investigational agent has passed the initial futility assessment (after approximately 150 participants per study arm are enrolled).

As a guideline, asymmetric boundaries will be provided to monitor the primary endpoint (time to sustained recovery) for overwhelming benefit or for harm. The trial of an investigational agent should be stopped for efficacy only if there is clear and convincing evidence of superiority of the agent versus the pooled control group with respect to the primary outcome, time to sustained recovery. For monitoring superiority, the Lan-DeMets spending function analogue of the O’Brien-Fleming boundaries will be used, with a 1-sided 0.025 level of significance over multiple looks. For computing the Lan-DeMets boundary, the information fraction at each interim analysis will be the observed total number of sustained recoveries divided by the planned number of sustained recoveries (N=843).

The monitoring boundary for harm is asymmetric, requiring less evidence to stop for harm than for superiority; a Haybittle-Peto boundary with 2.5 SD for the first 50 participants enrolled and 2.0 SD afterwards will used as a guideline for harm. With this approach, less evidence will be required for crossing a boundary for harm than for benefit.

At each full interim review after the first 169 participants have achieved sustained recovery (20% information time), the following will be provided:

- Signed square root of the value of the test statistic for Gray’s test with ρ=0, (“Z-value”) comparing the investigational agent versus the control group for the primary endpoint through Day 90, plotted over information time, and the asymmetric monitoring boundaries: the O’Brien-Fleming boundary with Lan-DeMets α-spending function for superiority (one-sided test with α=0.025), and the asymmetric, Haybittle-Peto boundary for harm described above.
  - **Comment:** Test statistics for the primary treatment comparison will be coded such that the value of the test statistic > 0 favors the investigational agent. Thus, in case of harm, the Haybittle-Peto boundary with 2 SD of the normalized test statistic, is crossed if the Z-value for the test statistic is below -2, irrespective of information time.

In addition to the current value of the test statistic, the corresponding values of the test statistic at the previous reviews will be plotted over information time, (1) as presented at the previous DSMB meetings, and (2) re-calculated with current data (using the cut-dates of the previous reports).

- History of the estimated rate ratios for time to sustained recovery with 95% CIs and p-values (by Fine-Gray’s method), and normalized test statistic values and p-values for Gray’s test at previous DSMB reviews, as presented, and recalculated with the current data (using the cut-date of the previous reports). The latter provides information on the influence of a possible time lag in the ascertainment of sustained recovery.

#### 9.4 Interim Monitoring for Futility

After investigational agents have passed the initial futility assessment (based on the pulmonary and pulmonary+ outcomes at Day 5, assessed for the first 300 participants), further futility analyses will be based on the primary outcome of *time to sustained recovery*.. The aim of these analyses will be to consider whether an investigational agent should be discontinued due to a low probability of achieving statistical significance for the primary endpoint of sustained recovery at the completion of the 90 day follow-up.

Conditional power calculations for time to sustained recovery will be presented under a range of scenarios. In the primary futility analysis, it will be assumed that the treatment effect for the future, as yet unobserved follow-up will be as hypothesized in the study design (RRR=1.25). As secondary analysis, the treatment effect for future follow-up will be assumed to be similar to the observed effect. Additional scenarios may be provided. Typical futility guidelines recommend stopping a trial when conditional power (assuming the originally hypothesized treatment effect for the future follow-up) is below 10%-15%.^11^

As a guideline, futility will first be assessed when 50% of the planned number of sustained recoveries have occurred, and a value of 15% will be suggested as a threshold for the conditional power. An additional assessment will take place at 75% of the events. Conditional power will be computed using Gray’s test with ρ=0, the competing risk analogue of the log-rank test.^12^

Decisions to terminate an agent for futility will include a broad assessment of the risk/benefit trade-off in addition to these guidelines.

### 10 Data Completeness and Study Conduct

According to the protocol, the pulmonary and pulmonary+ outcomes will be assessed on days 0-7; the decision rules for at the initial futility assessment are based on these outcomes on Day 5. The pulmonary outcome will also be assessed on Days 14 and 28. The primary outcome, “time to sustained recovery”, will be assessed through Day 90. Clinical data will be collected on Days 0-7, 14, 28, 60 and 90; mortality and re-hospitalizations will be assessed through 18 months. After hospital discharge, in-person visits are scheduled on Days 1, 3, 5, 28, and 90, when blood is collected (plasma and serum); other visits may be conducted by phone (Days 7, 14, 42, 60, and 75). The data collection schedule is included in Appendix D of this SAP.

Data completeness and study conduct reports will be provided by treatment group (for the closed report) and pooled across treatment groups (for the open report). Data summaries for the infusion of the investigational agent on Day 0 are described in Section 6; several of those reports are also relevant for monitoring study conduct and will be included in the open report or provided to study leadership, pooled across treatment groups.

The following data summaries will be provided to assess data completeness and study conduct:

- Number and percent of participants with protocol deviations, and type of protocol deviation
- Expected and observed number (% of expected) of participants who completed visits on Days 1-7, 14, 28, 42, 60, 75, and 90.
- Expected and observed number (% of expected) of participants with known outcome status for the pulmonary and the pulmonary+ outcomes on Day 5.
- Ascertainment of the primary outcome: Expected and observed number (% of expected) of participants with known status of “time to sustained recovery” at days 28, 60, and 90. To ascertain “sustained recovery”, several elements are required: vital status; the status of hospitalization; if discharged, the status of the residence (“home” versus other).
- Expected and observed number (% of expected) of participants with known vital status at days 5, 14, 28, 60 and 90, and at months 6, 12, and 18.
- Number and percent of participants who withdrew consent or were lost to follow-up (no contact and unknown vital status for 45+ days).
- If substantial numbers of participants are lost to follow-up (e.g., more than 10% of participants), Kaplan-Meier estimates for the cumulative proportion of participants who are lost to follow-up over time, by treatment group, will be provided (closed report only).
- Listing of participants who withdrew consent, including dates of randomization, pulmonary+ category at baseline, receipt of study treatment, date of withdrawal, and reason of withdrawal.
- Length of follow-up: Median, IQR, range
- Collection of specimens: Expected and observed number (% of expected) of participants with specimens collected as specified by the protocol, by visit.
- Expected and observed numbers of participants with local laboratory data at baseline and on Day 5.

A visit counts as “expected” if the visit window has closed or the data have been received.

### 11 SARS-CoV-2 Antibody Levels

SARS-CoV-2 antibody levels will be determined centrally, from stored plasma samples, and thus may not be available at interim analyses. If data are available, analyses will be included in interim reports.

Treatment groups will be compared for change in antibody profile, geometric mean titers (GMT) of antibodies and neutralizing antibody levels from baseline to Days 1, 3, 5, 28, and 90, using ANCOVA models applied to log-transformed antibody levels; usually, log_10_ is used for antibody titers.

Longitudinal models for the logarithm of antibody titers will be fit using GEE-based approaches to titers measured at baseline and days 1, 3, 5, 28 and 90; the interactions between time and group will be investigated. In addition, GMTs of the antibody levels will be summarized and compared between treatment groups at each of the days, using ANCOVA models for the log-transformed antibody titers.

The same approach will be used to examine neutralizing titers when such data are available.

Additional lab tests are currently being planned, including measurement of SARS-CoV-2 viral load and antigen levels. Analysis plans will be developed when more information is available.

### 12 Exploratory Analyses

#### 12.1 Associations Between the Pulmonary and Pulmonary+ Outcomes and Time to Sustained Recovery

At the early futility assessment when 300 participants (150 per study arm) are enrolled and have Day 5 data, estimated treatment differences in the pulmonary and pulmonary+ ordinal outcomes on Day 5 are used to identify promising investigational agents to continue to be studied with the clinical outcome of “time to sustained recovery”. In exploratory analyses, we will investigate whether the early pulmonary and pulmonary+ outcomes on Day 5 are an adequate predictor for time to sustained recovery, to re-evaluate our initial futility decision rule.

- Associations between the pulmonary and pulmonary+ ordinal outcomes at Day 5 with time to sustained recovery will be estimated using Cox proportional hazards models, pooled across treatment groups. To illustrate these associations, median time to sustained recovery will be estimated by category of the ordinal outcomes on Day 5, using Aalen-Johansen estimates of the cumulative incidence function.

Ideally, the goal would be to evaluate the extent to which treatment differences in the pulmonary outcomes on Day 5 predict treatment differences in time to sustained recovery through Day 90. A detailed analysis plan will be developed at a later time.

#### 12.2 Disease Progression Risk Score

A disease progression risk score will be developed, using pooled treatment groups with the following baseline predictors of the primary outcome (recovery): age, biological sex, duration of symptoms, ordinal outcome category at entry, and chronic health conditions. This risk score will be used for subgroup analyses for the primary outcome of time to sustained recovery, and time to mortality, to investigate if the treatment effect differs between subgroups at lower versus higher predicted disease progression risk.

A detailed analysis plan will be developed at a later time, but before analyses start.

### 13 Unblinding of Treatment Comparisons

For any investigational agent, trial results will be unblinded when the pre-specified number of primary endpoints is reached; results may be unblinded earlier upon the recommendation of the DSMB if the sponsor and study leadership concur. In this case, trial results for the investigational agent will be unblinded and reported with available data through 90 days of follow-up. After that, data collection will continue as outlined in the data collection plan; under protocol version 3.0, death and re-hospitalizations will be recorded through 18 months.

While the trial is ongoing, access to any data summaries by treatment group (investigational agent or control groups) will be restricted to the members of the DSMB, the DSMB’s Executive Secretary, and the unblinded statisticians.

When the trial for an investigational agent is concluded, data for the investigational agent and the corresponding pooled control group will be unblinded and provided to the study team.

The timing of the unblinding of data for one agent may require consideration, if:

- the control group is substantially shared with another agent for which the trial is still ongoing, **and**
- pooled data on treatment outcomes for the ongoing trial are available to investigators.

In this case, the need for a speedy unblinding has to be balanced with maintaining trial integrity for other agents in the platform trial, and the DSMB will be consulted as to the timing of the unblinding.

### 14 Distribution of Reports

- Open report: ACTIV-3 leadership team; DAIDS Medical Officer; selected NIAID staff; representatives of the companies; and all recipients of the unblinded closed report. After the DSMB meeting, the open report and the DSMB summary statement will be posted to the trial’s web site, open to all investigators.
- Closed report: DSMB members, Executive Secretary of the DSMB, unblinded statisticians.
- Web reports (accessible by all investigators and study staff):
  - Enrollment summaries by site and over time (updated daily)
  - Baseline characteristics
  - Selected summary measures on data quality and study conduct (pooled across treatment groups).
- Additionally, selected summary measures on study conduct will be provided to study leadership upon request (pooled across treatment groups).

Appendix A. Definition of the Pulmonary and Pulmonary+ ordered categorical outcomes

The Pulmonary categorical outcome is primarily defined based on oxygen requirements. The categories of the Pulmonary+ outcome are similar, except that categories 4 and 5 also capture selected extra-pulmonary complications, highlighted in red below.

| **Pulmonary outcome** | **Pulmonary+ outcome** |
| --- | --- |
| 1. Can independently undertake usual activities with minimal or no symptoms | 1. Can independently undertake usual activities with minimal or no symptoms |
| 2. Symptomatic and currently unable to independently undertake usual activities but no need of supplemental oxygen (or not above premorbid requirements) | 2. Symptomatic and currently unable to independently undertake usual activities but no need of supplemental oxygen (or not above premorbid requirements) |
| 3. Supplemental oxygen (<4 liters/min, or <4 liters/min above premorbid requirements) | 3. Supplemental oxygen (<4 liters/min, or <4 liters/min above premorbid requirements) |
| 4. Supplemental oxygen (≥4 liters/min, or ≥4 liters/min above premorbid requirements, but not high-flow oxygen) | 4. Supplemental oxygen (≥4 liters/min, or ≥4 liters/min above premorbid requirements, but not high-flow oxygen) or any of the following: stroke (NIH Stroke Scale [NIHSS] ≤14), meningitis, encephalitis, myelitis, myocardial infarction, myocarditis, pericarditis, new onset CHF NYHA class III or IV or worsening to class III or IV**,** arterial or deep venous thromboembolic events |
| 5. Non-invasive ventilation or high-flow oxygen | 5. Non-invasive ventilation or high-flow oxygen, or signs and symptoms of an acute stroke (NIHSS >14) |
| 6. Invasive ventilation, extracorporeal membrane oxygenation (ECMO), mechanical circulatory support, or new receipt of renal replacement therapy | 6. Invasive ventilation, ECMO, mechanical circulatory support, new receipt of renal replacement therapy, or vasopressor therapy |
| 7. Death | 7. Death |

The term “usual activities”, in categories 1 and 2 for both outcomes, refers to activities of daily living that the participant was able to undertake prior to the current illness.

Appendix B. Definition of Clinical Organ Failure and Serious Infection

According to the protocol, section 4.2.2., *clinical organ failure* is defined by development of any one or more of the following clinical events (see PIM for criteria for what constitutes each of these conditions):

a. Respiratory dysfunction:
   1. Respiratory failure defined as receipt of high flow nasal oxygen, non-invasive ventilation, invasive mechanical ventilation, or ECMO
b. Cardiac and vascular dysfunction:
   1. Myocardial infarction (MI)
   2. Myocarditis or pericarditis
   3. Congestive heart failure (CHF): new onset NYHA class III or IV, or worsening to class III or IV
   4. Hypotension requiring institution of vasopressor therapy
c. Renal dysfunction:
   1. New requirement for renal replacement therapy
d. Hepatic dysfunction:
   1. Hepatic decompensation
e. Neurological dysfunction
   1. Acute delirium
   2. Cerebrovascular event (stroke, cerebrovascular accident [CVA])
   3. Transient ischemic events (i.e., CVA symptomatology resolving <24 hrs)
   4. Encephalitis, meningitis or myelitis
f. Haematological dysfunction:
   1. Disseminated intravascular coagulation
   2. New arterial or venous thromboembolic events, including pulmonary embolism and deep vein thrombosis
   3. Major bleeding events (>2 units of blood within 24 hours, bleeding at a critical site [intracranial, intraspinal, intraocular, pericardial, intraarticular, intramuscular with compartment syndrome, or retroperitoneal], or fatal bleeding).

*Serious infection* is defined as:

g. Serious infection:
   1. Intercurrent, at least probable, documented serious disease caused by an infection *other than* SARS-CoV-2, requiring antimicrobial administration and care within an acute-care hospital.

Appendix C. Safety Data Collection

**Table C-1. Overview of Safety Data Collection** (protocol version 3, section 10).*

|  | Infusion +2 hrs | Days 0-7 | Day 14 | Day 28 | Day 90 | Months 6, 12, and 18 |
| --- | --- | --- | --- | --- | --- | --- |
| Infusion-related reactions and symptoms | X |  |  |  |  |  |
| Clinical AEs of any grade severity |  | X | X | X |  |  |
| Grade 3 and 4 clinical AEs from Day 7 through Day 28^1^ |  |  | X | X |  |  |
| Targeted laboratory abnormalities of any grade |  | X  (Day 5) |  |  |  |  |
| Hospital admissions and deaths |  | Collected through Month 18 | | | | |
| Targeted clinical events collected as study endpoints^2^ | Collected through Day 90 | | | | |  |
| SAEs not exempt from reporting (i.e., not considered a protocol specified exempt event)^2^ | Collected through Day 90 | | | | |  |
| Any SAE related to study intervention | Collected through Day 90 | | | | |  |
| Unanticipated problems | Collected through Day 90 | | | | |  |

^1^ Grade 3 and 4 clinical AEs on Days 1-7 are reported each day; those occurring between Days 8 and 14 are reported at the Day 14 visit, and those occurring between Days 15 and 28 are reported at the Day 28 visit.

^2^ Protocol-specified exempt serious events (PSEE); these events are listed below. See section 10.2.5 of the TICO study protocol and the PIM for information on PSEE.

* In protocol version 2.0, the data collection is identical, except that hospital re-admissions and deaths are collected through Month 12 only.

**Protocol-specified exempt events** (protocol section 10.2.5)

The following events are protocol-specified exempt events. They are **not** reported as AEs or SAEs, **unless** the investigator considered that there was a reasonable possibility that the study intervention (blinded investigational agent/ placebo or study-supplied SOC treatment) caused the event.

- Death
- Stroke
- Meningitis
- Encephalitis
- Myelitis
- Myocardial infarction
- Myocarditis
- Pericarditis
- New onset of worsening of CHF (NYHA class 3 or 4)
- Arterial or deep vein thromboembolic events
- Respiratory failure defined as receipt of high flow nasal oxygen, non-invasive ventilation, invasive mechanical ventilation or ECMO
- Hypotension requiring vasopressor therapy
- Renal dysfunction requiring renal replacement therapy
- Hepatic decompensation
- Neurologic dysfunction, including acute delirium and transient ischemic events
- Disseminated intravascular coagulation
- Major bleeding events
- Serious infections

Appendix D. Schedule of Assessments

**Table D-1. Schedule of Assessments** (protocol version 3.0, Appendix B).*

|  | **Screen or Day 0** | **Day 0** | **Follow-up Study Day; shaded columns denote in-person visits** | | | | | | | | | | | | | | | | |
| --- | --- | --- | --- | --- | --- | --- | --- | --- | --- | --- | --- | --- | --- | --- | --- | --- | --- | --- | --- |
| **Day** | **−1/0**^1^ | **0** | **1** | | **2** | **3** | **4** | **5** | **6** | **7** | **14** | **28** | **42** | **60** | **75** | **90** | **6M** | **12M** | **18M** |
| **Acceptable deviation from day** | **0** | **0** | **0** | | **0** | **0** | **0** | **0** | **0** | **+1** | **+2** | **+3** | **+3** | **+5** | **+5** | **+10** | **±14** | **±14** | **±14** |
| **ELIGIBILITY &  BASELINE DATA** |  |  |  | |  |  |  |  |  |  |  |  |  |  |  |  |  |  |  |
| Informed consent | X |  |  | |  |  |  |  |  |  |  |  |  |  |  |  |  |  |  |
| Baseline medical (incl. duration of COVID-19) and social history | X |  |  | |  |  |  |  |  |  |  |  |  |  |  |  |  |  |  |
| Baseline medications | X |  |  | |  |  |  |  |  |  |  |  |  |  |  |  |  |  |  |
| Symptom-directed physical exam by the clinical team | X |  |  | |  |  |  |  |  |  |  |  |  |  |  |  |  |  |  |
| Review SARS-CoV-2 test results | X |  |  | |  |  |  |  |  |  |  |  |  |  |  |  |  |  |  |
| Local laboratory testing | X |  |  | |  |  |  | X |  |  |  |  |  |  |  |  |  |  |  |
| Urine pregnancy test or other documentation of pregnancy status | X |  |  | |  |  |  |  |  |  |  |  |  |  |  |  |  |  |  |
| **STUDY INTERVENTION** |  |  |  | |  |  |  |  |  |  |  |  |  |  |  |  |  |  |  |
| Randomization |  | X |  | |  |  |  |  |  |  |  |  |  |  |  |  |  |  |  |
| Study Drug/Placebo Administration |  | X |  | |  |  |  |  |  |  |  |  |  |  |  |  |  |  |  |
| Assess infusion completion and adverse reactions |  | X |  | |  |  |  |  |  |  |  |  |  |  |  |  |  |  |  |
| **STUDY PROCEDURES** |  |  |  | |  |  |  |  |  |  |  |  |  |  |  |  |  |  |  |
| Clinical assessment for pulmonary ordinal outcome | X | X | X | | X | X | X | X | X | X | X | X |  |  |  |  |  |  |  |
| Clinical assessment for pulmonary+ ordinal outcome | X | X | X | | X | X | X | X | X | X |  |  |  |  |  |  |  |  |  |
| Vital signs for NEW score assessment | X |  |  | |  |  |  |  |  |  |  |  |  |  |  |  |  |  |  |
| Respiratory function scale assessment | X |  |  | |  |  |  |  |  |  |  |  |  |  |  |  |  |  |  |
| Hospitalization status |  |  |  | |  | X |  | X |  | X | X | X |  | X |  | X | X | X | X |
| Changes in residence/facility |  |  |  | |  |  |  |  |  |  | X | X | X | X | X | X |  |  |  |
| Interim medical history |  |  |  | |  |  |  |  |  | X | X | X |  | X |  | X |  |  |  |
| Interim medications |  |  |  | |  |  |  | X |  |  |  | X |  |  |  |  |  |  |  |
| Clinical AEs of any grade, on days indicated |  | X | X | | X | X | X | X | X | X | X | X |  |  |  |  |  |  |  |
| Clinical AEs reaching grade 3 or 4 severity through Day 28 |  |  |  | |  |  |  |  |  |  | X | X |  |  |  |  |  |  |  |
| Research sample storage (plasma and serum)^2^ |  | X | X | |  | X |  | X |  |  |  | X |  |  |  | X |  |  |  |
| Mid*turbinate* swab for central SARS-CoV-2 viral load testing^2^ |  | X |  | |  |  |  |  |  |  |  |  |  |  |  |  |  |  |  |
| SAEs and unanticipated problems |  |  | Report as they occur | | | | | | | | | | | | | |  |  |  |
| Deaths |  |  | Report as they occur | | | | | | | | | | | | | | | | |
| Hospitalization Summary |  |  | Report upon hospital discharge | | | | | | | | | | | | | |  |  |  |
| Hospital Readmissions |  |  | | Report upon hospital discharge | | | | | | | | | | | | | | | |

^1^ Screening must be performed within 24 hours of randomization.

^2^ Blood draw and swab collection in some cases can be obtained after randomization but before the infusion. If it is not possible to do an in-person on Day 3 or Day 5, the blood draws may be done one day earlier or one day later (but the participant should be telephoned to record the clinical data on the indicated study day).

* In protocol version 2.0, the data collection is identical, except that hospital re-admissions and deaths are collected through Month 12 only.

Appendix E. List of Acronyms

ACTIV	Accelerating COVID-19 Therapeutic Interventions and Vaccines

ACTT	[Adaptive COVID-19 Treatment Trial](https://www.niaid.nih.gov/news-events/nih-clinical-trial-remdesivir-treat-covid-19-begins)

ADE	Antibody-dependent enhancement

AE	Adverse event

ARDS	Acute respiratory distress syndrome

CHF	Congestive heart failure

CHF	Coronary heart failure

CI	Confidence interval

CIF	Cumulative incidence curve

CMH	Cochran-Mantel-Haenszel [test]

COVID-19	Coronavirus-Induced Disease 2019

CVA	Cerebrovascular accident

DSMB	Data and Safety Monitoring Board

ECMO	Extracorporeal membrane oxygenation

EU	European Union

FDA	Food and Drug Administration (US)

GCP	Good Clinical Practice

GDPR	General Data Protection Regulation

GEE	Generalized estimating equations

GMT	Geometric mean titer

HR	Hazard ratio

ICC	International Coordinating Center

ICH	International Council for Harmonisation of Technical Requirements for Pharmaceuticals for Human Use

ICU	Intensive care unit

IgG	Immunoglobulin G

IL-6	Interleukin 6

INSIGHT	International Network for Strategic Initiatives in Global HIV Trials

IQR	Interquartile range

IRB	Institutional Review Board

ITT	Intention-to-treat

IV	Intravenous

mAb	Monoclonal antibody

MedDRA	Medical Dictionary for Regulatory Activities

MI	Myocardial infarction

mITT	modified intention-to-treat

mL	Milliliter

NEW	National Early Warning [score]

NIAID	National Institute of Allergy and Infectious Diseases, NIH (US)

NIH	National Institutes of Health (US)

NIHSS	National Institutes of Health Stroke Scale/Score

NYHA	New York Heart Association

nMAb	Neutralizing Monoclonal Antibodies

OR	Odds ratio

PCR	Polymerase chain reaction

PIM	Protocol Instruction Manual

PT	Preferred term

PSEE	Protocol-specified exempt (serious) events

RNA	Ribonucleic acid

RR	Rate ratio

RRR	Recovery rate ratio

SAE	Serious adverse event

SARS-CoV-1	Severe acute respiratory syndrome coronavirus 1

SARS-CoV-2	Severe acute respiratory syndrome coronavirus 2

SAP	Statistical analysis plan

SOC	Standard of care

SUSAR	Suspected unexpected serious adverse reaction

TOC	Trial oversight committee

UMN	University of Minnesota

UP	Unanticipated problem

U.S.	United States of America

WHO	World Health Organization

1. The Medical Dictionary for Regulatory Activities terminology is the international medical terminology developed under the auspices of the International Conference on Harmonization of Technical Requirements for Registration of Pharmaceuticals for Human Use (ICH). MedDRA® is a registered trademark of the International Federation of Pharmaceutical Manufacturers and Associations (IFPMA)
